# Plasma Neurofilament Light Chain and Glial Fibrillary Acidic Protein in Psychiatric Disorders: A Large-Scale Normative Modeling Study

**DOI:** 10.64898/2026.04.08.26350391

**Authors:** Anna M. Jacobsen, Boris B. Quednow, Francesco Bavato

## Abstract

**Importance:** Blood neurofilament light chain (NfL) and glial fibrillary acidic protein (GFAP) are entering clinical use in neurology as markers of neuroaxonal and astrocytic injury, but their utility in psychiatry is unclear.

**Objective:** To determine whether psychiatric diagnoses are associated with altered plasma NfL and GFAP levels.

**Design, Setting, and Participants:** This population-based study examined plasma NfL and GFAP among 47,495 participants from the UK Biobank (54.0% female; 93.5% White; mean [SD] age 56.8 [8.2] years) who provided blood samples and sociodemographic and clinical data between 2006 and 2010. Normative modeling was applied to assess associations between 7 lifetime psychiatric diagnostic categories and deviations from expected NfL and GFAP levels, while accounting for neurological diagnoses, cardiometabolic burden, and substance use. Data were analyzed between July 2025 and March 2026.

**Main Outcomes and Measures:** Deviations in plasma NfL and GFAP levels from normative predictions.

**Results:** Relative to the reference population, plasma NfL levels were higher among individuals with bipolar disorder (d=0.20; 95% CI, 0.03–0.37; p=0.03), recurrent depressive disorder (d=0.23; 95% CI, 0.07–0.38; p=0.009), and depressive episodes (d=0.06; 95% CI, 0.02–0.10; p=0.01), lower among individuals with anxiety disorders (d=−0.07; 95% CI, −0.12 to −0.02; p=0.008), but did not differ in schizophrenia spectrum, stress-related, or other psychiatric disorders. Plasma GFAP levels were not elevated in any psychiatric disorders. Variability in NfL levels was greater among individuals with schizophrenia spectrum disorders (variance ratio [VR]=1.30; p=0.005), depressive episodes (VR=1.06; p=0.006), and anxiety disorders (VR=1.08; p=0.005). Variability in GFAP levels was increased only in anxiety disorders (VR=1.08; p=0.01). Plasma NfL levels exceeding percentile-based normative thresholds were more common among individuals with schizophrenia spectrum disorders, bipolar disorder, recurrent depressive disorder, and depressive episodes. Neurological diagnoses, cardiometabolic burden, and substance use were associated with plasma NfL and GFAP levels.

**Conclusions and Relevance:** This study provides population-level evidence of plasma NfL elevation in bipolar and depressive disorders and increased variability in schizophrenia spectrum, bipolar and depressive disorders, supporting its potential as a biomarker in psychiatry and informing its ongoing neurological applications. Plasma GFAP levels, in contrast, were largely unaltered across psychiatric disorders.

**Key Points:** *Question:* Are plasma neurofilament light chain (NfL) and glial fibrillary acidic protein (GFAP) levels altered in psychiatric disorders?

*Findings:* In this cohort study including 47,495 individuals, normative modeling revealed that plasma NfL levels were elevated in bipolar and depressive disorders, whereas plasma GFAP levels were not elevated in any psychiatric disorder. Plasma NfL levels also showed higher variability in schizophrenia spectrum, bipolar, and depressive disorders.

*Meaning:* Plasma NfL shows distinct alterations in schizophrenia spectrum and affective disorders, supporting its further investigation as a biomarker in clinical psychiatry and highlighting the need to consider psychiatric comorbidity in neurological applications.

## Introduction

Unlike other medical fields, psychiatry lacks biologically informed tools to stratify patients or monitor treatments based on brain-related outcomes.^1^ This represents a critical barrier to the implementation of precision medicine in psychiatry.^2^

Neurofilament light chain (NfL) and glial fibrillary acidic protein (GFAP) are cytoskeletal proteins exclusively expressed in neurons and astrocytes, respectively, that are detectable in peripheral blood. Blood NfL and GFAP have recently been established as reliable markers of brain aging and injury, with increased levels reflecting structural brain changes in neuroinflammatory, cerebrovascular, traumatic, and neurodegenerative conditions.^3, 4^ Both markers are increasingly used in neurological practice for diagnostic evaluation, prognostic assessment, and treatment monitoring.^5^ However, their utility in psychiatry remains unclear.^6^

Previous investigations of blood NfL and GFAP in primary psychiatric disorders have yielded mixed results, potentially due to small sample sizes and heterogeneous consideration of relevant demographic and clinical confounders.^6^ Several studies have shown altered blood NfL levels among samples or subgroups of individuals with depression, schizophrenia, and bipolar disorder, but other investigations among the same clinical populations have shown null or mixed findings.^7–11^ Few studies assessing blood GFAP levels in psychiatric disorders have shown similarly promising yet inconclusive findings.^12–14^ Large-scale evidence for NfL and GFAP alterations in psychiatry is still limited, and filling this gap is a necessary first step to clarify their potential clinical utility.

Here we leveraged the UK Biobank dataset to examine plasma NfL and GFAP levels across psychiatric disorders. We generated normative models to predict plasma NfL and GFAP levels, then assessed how deviations from normative values were associated with lifetime diagnoses of schizophrenia spectrum disorders, bipolar disorder, recurrent depressive disorder, single depressive episodes, anxiety disorders, stress-related disorders, and other psychiatric disorders when accounting for cardiometabolic burden and other relevant demographic and clinical factors.^15^ We further quantified analogous deviations in neurological disorders and included neurological comorbidities in all models. Additionally, we examined group differences in variability and estimated the odds of NfL and GFAP levels exceeding normative percentile-based thresholds as indices of biological heterogeneity in psychiatric populations. We hypothesized that primary psychiatric disorders would be associated with elevated plasma NfL and GFAP levels and increased variability relative to normative values after adjustment for neurological comorbidities and demographic and clinical factors.

## Methods

### Cohort Description

Participants aged 40 to 70 years were recruited between 2006 and 2010 from 22 sites as part of the UK Biobank, an ongoing population-based cohort study in the United Kingdom (application 853656).^16^ The North West Multicenter Research Ethics Committee approved the study, and all participants provided written informed consent. Participants completed sociodemographic and lifestyle questionnaires, granted access to healthcare records, underwent physical examination, and provided biological samples. Corresponding UK Biobank variables are listed in Supplementary Table 1. Analyses were restricted to participants with available NfL and GFAP plasma proteomics data who completed all relevant questionnaires (N=47,995; Supplementary Figure 1).

Psychiatric diagnostic categories were defined using lifetime ICD10 diagnoses from participant healthcare records (Supplementary Table 2). Psychiatric categories included schizophrenia spectrum disorders (F20–29), bipolar disorder (F30–31), recurrent depressive disorder (F33), a depressive episode (F32), anxiety (F40–41), and stress-related disorders (F43). Neurological categories included neurodegenerative (F00–03, G10, G12–13, G20, G23, G25, G30–32), neuroinflammatory (G09, G35–37), neuromuscular (G11, G70–73), cerebrovascular (G45–46, I60–69), peripheral neuropathic (G60–64), and epilepsy (G40–41) disorders. Less prevalent diagnoses without a priori hypotheses were grouped as “other psychiatric” (F34, F38–39, F42, F44, F48, F99) or “other neurological” (G80–83, G91–99) disorders. The reference population consisted of participants without lifetime psychiatric or neurological diagnoses. Individuals with infectious nervous system diseases (G00–08, G14, I02) before blood sampling or lifetime neurodevelopmental diagnoses (F70–72, F78–84, F89–90) were excluded (Supplementary Figure 1). Healthcare records were last updated in December 2025.

### Blood Markers

Blood samples were collected between March 2006 and November 2010 according to UK Biobank protocol.^16^ EDTA plasma aliquots were stored at –80°C at the UK Biobank coordinating center (Stockport, England, UK). As part of the Pharma Proteomics Project, plasma NfL and GFAP were quantified via proximity extension assay between April 2021 and June 2022 (Olink Analysis Services; Uppsala, Sweden).^17^ This analysis generated normalized protein levels that represent relative plasma NfL and GFAP abundance.

### Statistical Analysis

We generated normative models for plasma NfL and GFAP levels among individuals with no lifetime incidence of psychiatric or neurological disorders using Bayesian linear regression from *pcntoolkit* (v1.1.2; Supplementary Figure 2). Age and body mass index (BMI) were included as covariates with sex as a batch effect due to the well-established associations of these factors with both markers.^3, 4^ Blood marker levels were transformed with sin-hyperbolic arcsin warping from *pcntoolkit* and BMI was log-transformed to ensure normal distributions. Nonlinear contributions of continuous covariates were modeled with B-spline basis function (3 degrees of freedom).^18^ The normative models explained 30% of plasma NfL variance and 15% of plasma GFAP variance. Diagnostic metrics indicated robust predictive performance (Supplementary Table 3). Of n=28,539 participants with no lifetime incidence of psychiatric or neurological disorders, n=5708 randomly assigned to the testing dataset were considered the reference population, while n=22,831 participants comprising the normative modeling training dataset were not included in further analyses to avoid circularity. Normative blood marker levels were predicted for all participants in reference, psychiatric, and neurological populations from their age, sex, and BMI. Deviation scores (z-scores) were computed to quantify individual divergence from expected values.

To evaluate associations between psychiatric and neurological diagnoses and NfL and GFAP z-scores, we used linear mixed effects models (*statsmodels* v0.14.6). Participants could belong to multiple categories to quantify differential diagnostic contributions. No overlap was allowed between the bipolar disorder, recurrent depressive disorder, and depressive episode categories due to their definitional mutual exclusion. Linear models were adjusted for cardiometabolic burden, self-reported tobacco smoker status, and self-reported alcohol use frequency (Supplementary Figure 3) given their known associations with blood NfL and GFAP levels.^4, 19, 20^ Covariates were examined individually, then jointly to assess their independent and combined contributions. Cardiometabolic burden was quantified based on lifetime ICD10 diagnoses related to heart disease, hypertension, diabetes, and reduced renal function, with one point assigned for each category with at least one diagnosis present (Supplementary Table 4). To adjust for multiple comparisons, we applied Benjamini-Hochberg false discovery control to linear model p-values for all 14 diagnostic categories (*scipy* v1.15.3).

Adjusted z-scores were derived for each psychiatric diagnostic category using the corresponding linear model coefficient and individual participant residuals (Supplementary Figure 4). We analyzed variance differences in adjusted z-scores between psychiatric and reference populations with Levene’s test and calculated variance ratios between psychiatric and reference populations (*scipy* v1.15.3).

Consistent with established approaches for identifying extreme blood marker deviations, we also quantified the percentage of individuals in psychiatric populations with z-scores exceeding normative 80^th^, 90^th^, and 95^th^ percentiles.^3^ To estimate the probability that each psychiatric population contained participants with z-scores exceeding these cutoffs, we employed logistic regression adjusted for psychiatric and neurological comorbidities, cardiometabolic burden, tobacco use, and alcohol use (*statsmodels* v0.14.6). Odds ratios for psychiatric diagnoses were generated by exponentiating their corresponding logistic regression coefficients.

## Results

### Cohort Characteristics

Our cohort of 47,495 participants included 309 individuals with lifetime diagnoses of schizophrenia spectrum disorders, 283 with bipolar disorder, 326 with recurrent depressive disorder, 5994 with a depressive episode, 4728 with anxiety disorders, 1496 with stress-related disorders, and 815 with other psychiatric disorders. The reference population included 5708 individuals without lifetime incidence of psychiatric or neurological disorders. Psychiatric and reference population characteristics are reported in Table 1. Our cohort also included 3109 individuals with neurodegenerative disorders, 489 with neuroinflammatory disorders, 213 with neuromuscular disorders, 3996 with cerebrovascular disorders, 887 with peripheral neuropathic disorders, 940 with epilepsy, and 2284 with other neurological disorders. Neurological population characteristics are reported in Supplementary Table 5. Psychiatric and neurological populations differed from the reference population in most demographic and lifestyle characteristics we considered (Supplementary Table 6).

**Table 1.**
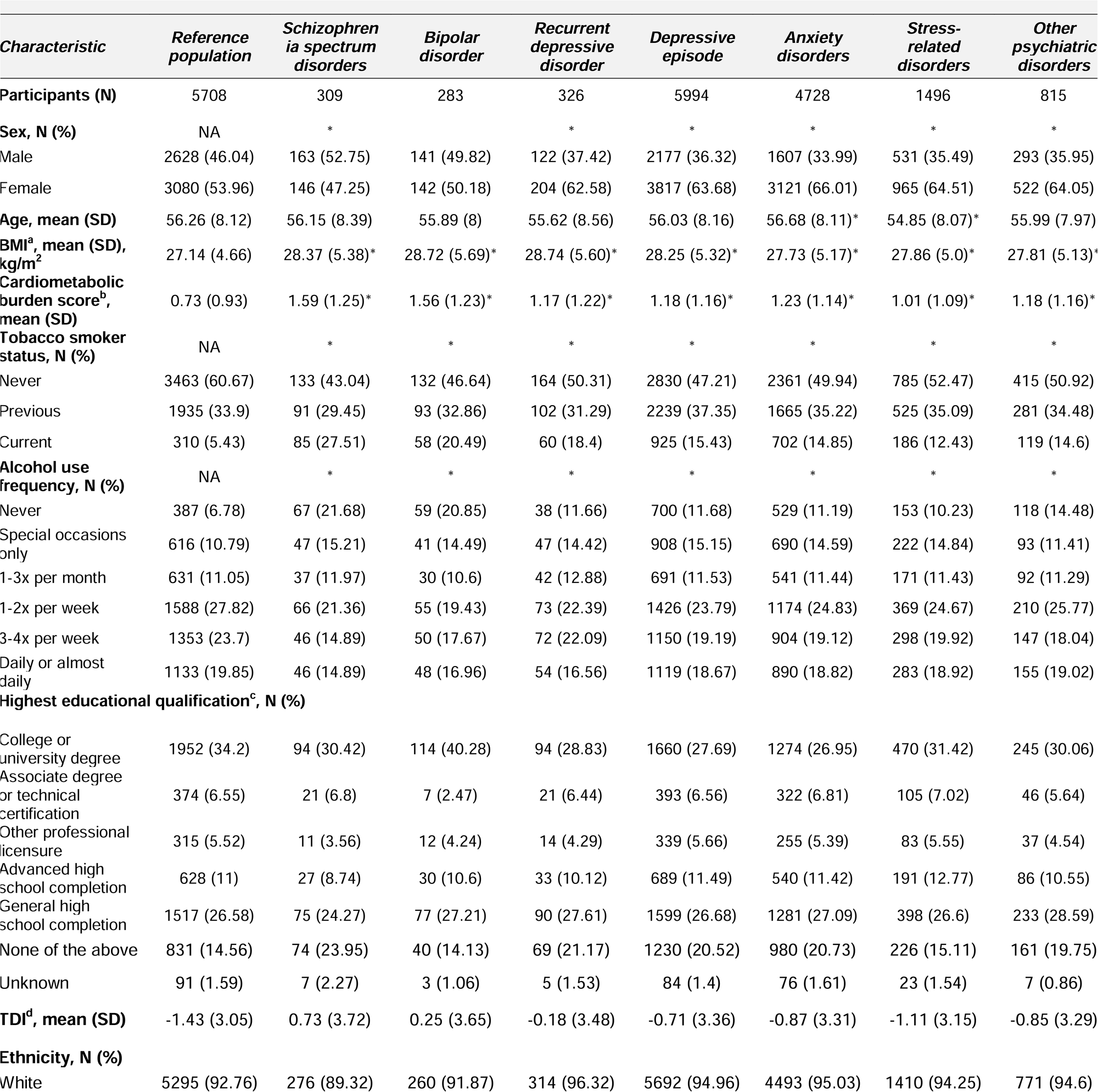
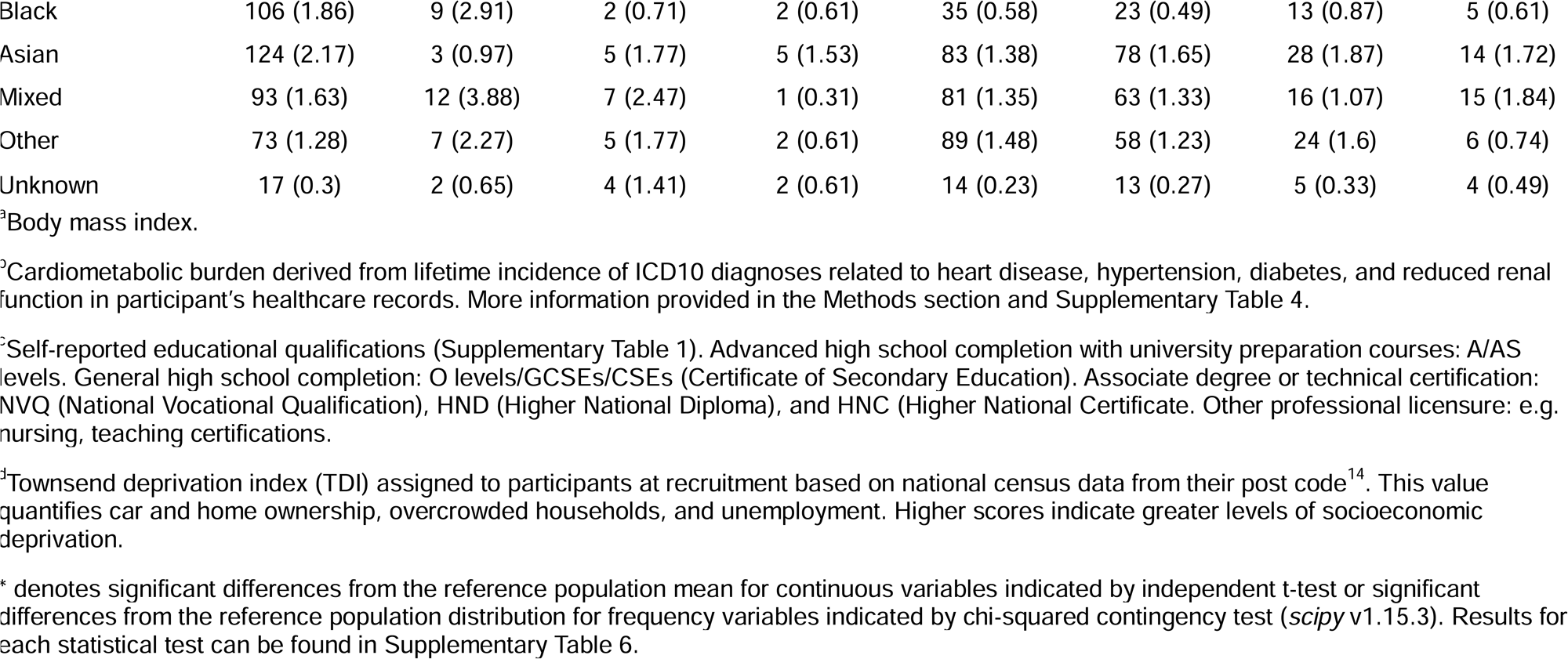
Demographic and Clinical Characteristics of Psychiatric and Reference Populations.

### NfL and GFAP Associations with Psychiatric and Neurological Diagnoses

We generated NfL and GFAP z-scores for psychiatric, neurological, and reference populations that describe deviation from normative predictions of plasma NfL and GFAP levels given age, sex, and BMI. When accounting for psychiatric and neurological comorbidities, cardiometabolic burden, tobacco use, and alcohol use (Figure 1), elevated NfL z-scores were associated with lifetime diagnoses of bipolar (d=0.20; 95% CI, 0.03-0.37; p=0.03) and recurrent depressive disorders (d=0.23; 95% CI, 0.07-0.38; p=0.009) as well as a depressive episode (d=0.06; 95% CI, 0.02-0.10; p=0.01). Diagnoses of schizophrenia spectrum (d=-0.05; 95% CI, – 0.21 to 0.11; p=0.56), stress-related (d=0.01; 95% CI, –0.06 to 0.09; p=0.70), and other psychiatric disorders (d=-0.04; 95% CI, –0.14 to 0.06; p=0.45) were not significantly associated with NfL z-scores, while anxiety disorder diagnoses were associated with lower NfL z-scores (d=-0.07; 95% CI, –0.12 to –0.02; p=0.008). All neurological diagnostic categories were associated with increased NfL z-scores (neurodegenerative disorders: d=0.28; 95% CI, 0.22-0.33; p<0.001; neuroinflammatory disorders: d=0.46; 95% CI, 0.33-0.59; p<0.001; neuromuscular disorders: d=0.21; 95% CI, 0.02-0.40; p=0.04; cerebrovascular disorders: d=0.08; 95% CI, 0.02-0.13; p=0.01; peripheral neuropathic disorders: d=0.54; 95% CI, 0.44-0.63; p<0.001; epilepsy: d=0.26; 95% CI, 0.17-0.36; p<0.001; other neurological disorders: d=0.09; 95% CI, 0.03-0.16; p=0.008).

**Figure 1.**
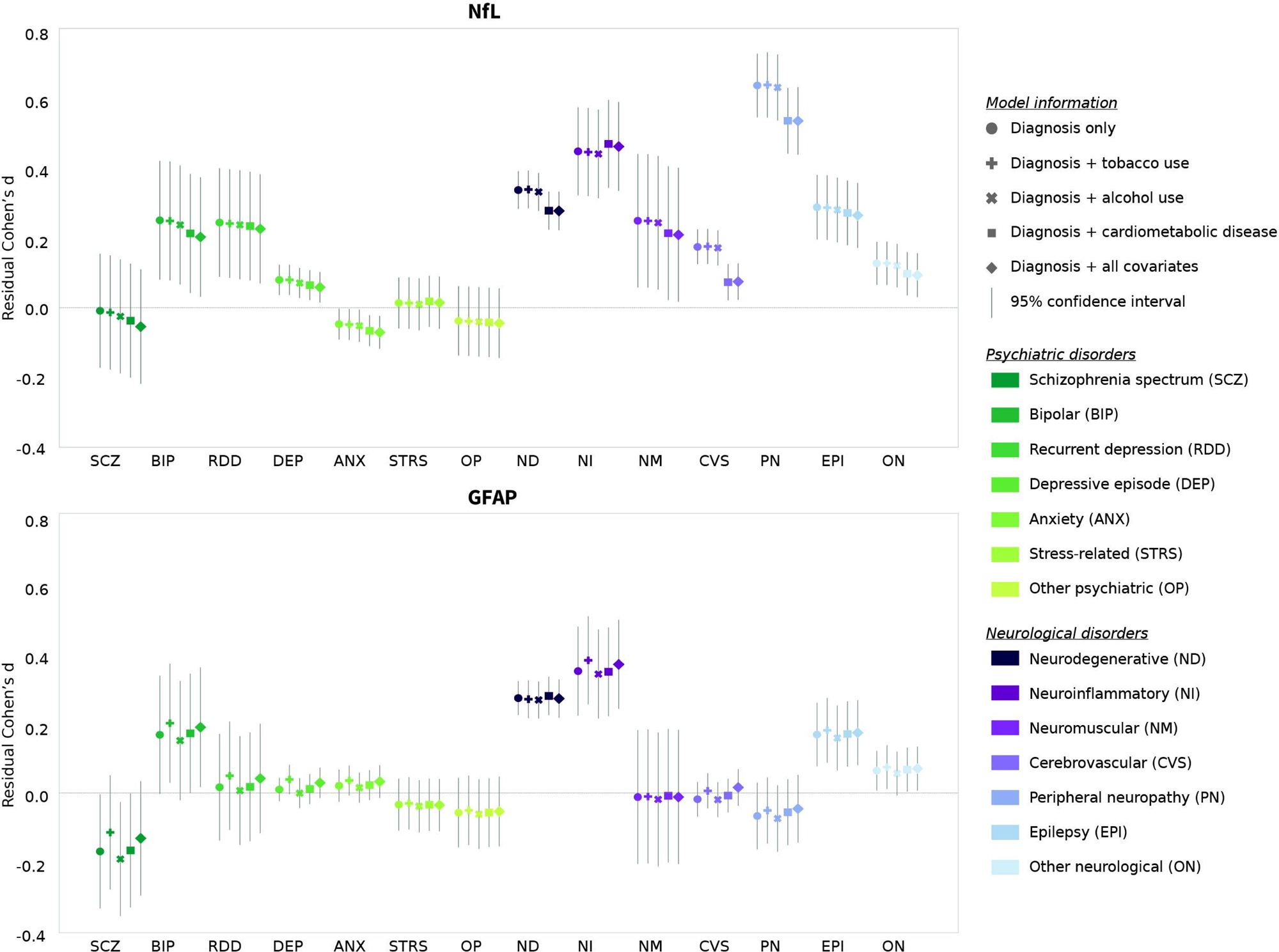
Plasma NfL and GFAP Z-score Associations with Lifetime Incidence of Psychiatric and Neurological Disorders. Residual Cohen’s d (effect size) with 95% confidence intervals for association between lifetime incidence of psychiatric and neurological diagnoses and NfL and GFAP z-scores adjusted for psychiatric and neurological comorbidities and, separately, tobacco use, alcohol use, and cardiometabolic burden, as well as adjusted for all covariates together.

After the same adjustments, elevated GFAP z-scores were not associated with any psychiatric disorder (schizophrenia spectrum disorders: d=-0.13; 95% CI, –0.29-0.03; p=0.28; bipolar disorder: d=0.19; 95% CI, 0.02-0.36; p=0.09; recurrent depressive disorder: d=0.04; 95% CI, –0.11 to 0.20; p=0.65; depressive episode: d=0.03; 95% CI, –0.01 to 0.07; p=0.33; anxiety disorders: d=0.03; 95% CI, –0.01 to 0.08; p=0.33; stress-related disorders: d=-0.03; 95% CI, –0.11 to 0.04; p=0.46; other psychiatric disorders: d=-0.05; 95% CI, –0.15 to 0.05; p=0.46). Neurodegenerative disorders (d=0.27; 95% CI, 0.22-0.32; p<0.001), neuroinflammatory disorders (d=0.37; 95% CI, 0.24-0.50; p<0.001), and epilepsy (d=0.17; 95% CI, 0.08-0.27; p=0.001) were significantly associated with elevated with elevated GFAP z-scores, but not neuromuscular (d=-0.01; 95% CI, –0.20 to 0.18; p=0.91), cerebrovascular (d=0.02; 95% CI, – 0.04 to 0.07; p=0.64), peripheral neuropathic disorders (d=-0.05; 95% CI, –0.14 to 0.05; p=0.46), or other neurological disorders (d=0.07; 95% CI, 0.007-0.13; p=0.09).

Cardiometabolic burden, tobacco use, and alcohol use were significantly associated with NfL and GFAP z-scores in all models. Their inclusion as covariates altered the magnitude and significance of associations between psychiatric and neurological diagnoses and NfL and GFAP z-scores (Figure 1; Supplementary Tables 7-9).

### NfL and GFAP Variability Among Psychiatric Populations

We compared variability in NfL and GFAP z-scores between psychiatric and reference populations, adjusted for psychiatric and neurological comorbidities, cardiometabolic burden, tobacco use, and alcohol use. Higher variability in NfL z-scores occurred among individuals with lifetime diagnoses of schizophrenia spectrum disorders (variance ratio (VR)=1.30; F_1, 6016_=8.07; p=0.005), a depressive episode (VR=1.06; F_1, 11701_=7.60; p=0.006), and anxiety disorders (VR=1.08; F_1, 10435_=7.91; p=0.005), but not individuals with bipolar (VR=1.10; F_1, 5990_=2.10; p=0.15), recurrent depressive (VR=1.18; F_1, 6033_=2.03; p=0.15), stress-related (VR=1.0; F_1, 7203_=0.25; p=0.62), or other psychiatric disorders (VR=1.02; F_1, 6522_=0.07; p=0.78).

For GFAP z-scores, higher variability occurred among individuals with anxiety disorders (VR=1.08; F_1, 10435_=6.44; p=0.01), but not among individuals with schizophrenia spectrum disorders (VR=1.19; F_1, 6016_=2.34; p=0.13), bipolar disorder (VR=1.01; F_1, 5990_=0.007; p=0.94), recurrent depressive disorder (VR=1.0; F_1, 6033_=0.20; p=0.66), a depressive episode (VR=1.05; F_1, 11701_=2.16; p=0.14), stress-related disorders (VR=1.01; F_1, 7203_=0.60; p=0.44), or other psychiatric disorders (VR=1.02; F_1, 6522_=0.02; p=0.90).

### NfL and GFAP in Psychiatric Populations Compared to Normative Thresholds

We quantified the proportion of and estimated odds ratios (ORs) for individuals in psychiatric populations with NfL and GFAP z-scores exceeding normative 80th, 90th, and 95th percentile thresholds, adjusted for psychiatric and neurological comorbidities, cardiometabolic burden, tobacco use, and alcohol use (Figure 2). Among individuals with schizophrenia spectrum disorders, odds of NfL z-scores exceeding normative thresholds were increased for the 95^th^ percentile (11.65%; OR=1.68; 95% CI, 1.14–2.46) but not for the 80^th^ (27.18%; OR=0.92; 95% CI, 0.70–1.20) or 90^th^ (16.18%; OR=1.10; 95% CI, 0.79–1.53) percentiles. Individuals with bipolar disorder showed increased odds of NfL z-scores exceeding the 80^th^ (30.39%; OR=1.33; 95% CI, 1.01–1.74) and 90^th^ (18.02%; OR=1.59; 95% CI, 1.14–2.21) percentiles but not the 95^th^ (9.54%; OR=1.44; 95% CI, 0.93–2.24). Among individuals with recurrent depressive disorder, NfL z-scores were more likely to exceed the 80^th^ (29.45%; OR=1.47; 95% CI, 1.15–1.89), 90^th^ (14.11%; OR=1.48; 95% CI, 1.07–2.05), and 95^th^ (7.67%; OR=1.62; 95% CI, 1.06–2.48) percentiles. Increased odds of higher NfL z-scores were also observed among individuals with a depressive episode across all percentiles (80^th^: 24.91%; OR=1.14; 95% CI, 1.06–1.23; 90^th^: 11.80%; OR=1.17; 95% CI, 1.06–1.29; 95^th^: 5.97%; OR=1.21; 95% CI, 1.06–1.39). In contrast, anxiety disorders were associated with lower odds of exceeding the 90^th^ (10.32%; OR=0.86; 95% CI, 0.77–0.96) and 95^th^ (4.76%; OR=0.77; 95% CI, 0.66–0.90) percentiles, with no difference at the 80^th^ percentile (23.12%; OR=0.93; 95% CI, 0.86–1.01). No significant associations were observed for stress-related disorders (80^th^: 23.20%; OR=1.05; 95% CI, 0.92–1.19; 90^th^: 10.36%; OR=1.02; 95% CI, 0.85–1.21; 95^th^: 4.75%; OR=0.95; 95% CI, 0.74–1.22) or other psychiatric disorders (80^th^: 22.09%; OR=0.88; 95% CI, 0.74–1.05; 90^th^: 10.43%; OR=0.90; 95% CI, 0.71–1.14; 95^th^: 4.79%; OR=0.83; 95% CI, 0.59–1.16). GFAP z-scores were not more likely to exceed normative percentile thresholds in any psychiatric populations (Supplementary Table 10).

**Figure 2.**
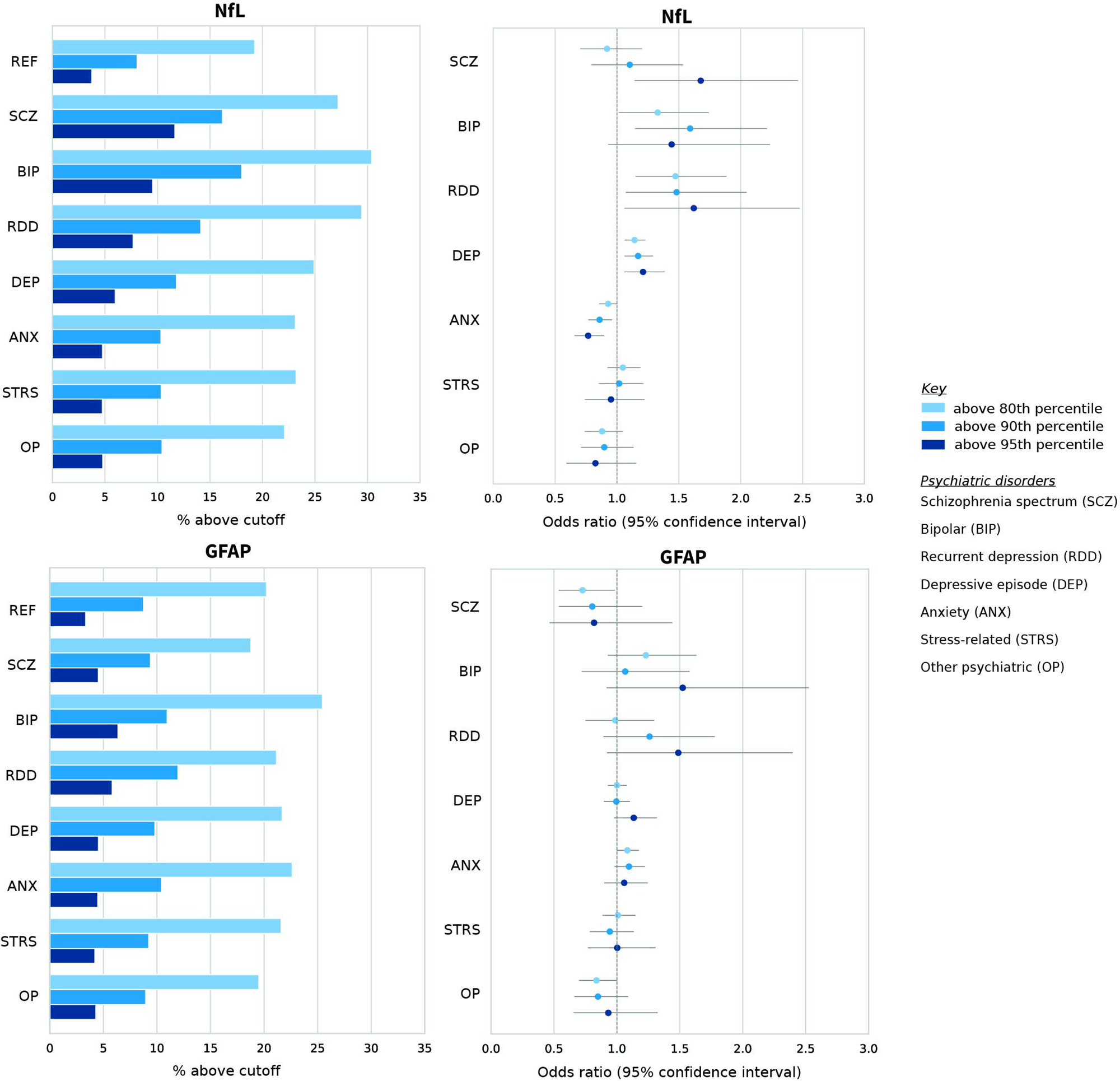
Plasma NfL and GFAP Z-scores Above Normative Thresholds in Psychiatric Populations. Left: Percentage of NfL and GFAP z-scores in psychiatric and reference populations above normative 80th, 90th, and 95th percentiles. Right: Odds ratios describing likelihood that psychiatric populations contain individuals with NfL and GFAP z-scores above normative 80th, 90th, and 95th percentiles based on logistic regression adjusted for psychiatric and neurological comorbidities, cardiometabolic burden, tobacco use, and alcohol use.

## Discussion

In this large population-based study, plasma NfL levels were elevated in affective disorders and showed increased variability in both schizophrenia spectrum and affective disorders, whereas plasma GFAP alterations were limited to increased variability in anxiety disorders.

Plasma NfL elevations observed among individuals with affective disorders align with prior research and with neuroimaging evidence of reduced structural integrity and accelerated brain aging in these disorders.^7,21,22^ Anxiety disorders, in contrast, were associated with slightly lower plasma NfL levels. Speculative explanations include compensatory changes in neuroaxonal turnover or medication effects. Overall, the small NfL alterations in affective disorders do not support its use for individual diagnosis, similar to neurological applications where blood NfL is primarily used to monitor disease activity.^23^ Nevertheless, NfL elevations in bipolar and recurrent depressive disorders overlapped with those reported in several neurological populations, suggesting clinically relevant alterations. Plasma NfL may therefore help identify biologically defined subgroups, predict treatments, monitor disease trajectories, and evaluate neuroprotective interventions in affective disorders, although longitudinal studies with detailed clinical characterization are needed.^6^

In schizophrenia spectrum disorder, consistent with most previous studies, we did not observe elevated plasma NfL levels.^7^ Early-stage brain pathology in schizophrenia detectable with neuroimaging may not be reflected in plasma NfL, a marker of active neuroaxonal injury.^21,24^ However, we observed greater variance and a higher prevalence of individuals with plasma NfL exceeding normative thresholds in this population, also consistent with previous findings.^7^ This pattern aligns with current hypotheses of biological heterogeneity within psychotic disorders.^25, 26^ Prior work has pointed to the existence of subgroups with more pronounced neuroaxonal alterations that are potentially linked to reduced treatment response, but further investigation is needed to establish the clinical and mechanistic correlates of this variability.^7, 27, 28^ Given the proposed role of blood NfL in distinguishing primary psychiatric disorders from secondary psychosis in neuroinflammatory conditions, extreme deviations warrant further investigation.^29^

Evidence for GFAP alterations among individuals with psychiatric disorders in this sample was limited. Increased variability in plasma GFAP was observed only in anxiety disorders. Limited associations between psychiatric diagnoses and plasma GFAP compared to associations with plasma NfL likely reflect distinct biological properties of the two markers. Blood GFAP may capture transient astrocytic responses that are less detectable at the population level, particularly when clinical populations are defined by lifetime diagnoses.^4, 30^

Because NfL and GFAP are increasingly used in neurology, these findings have implications beyond psychiatry. The association of schizophrenia spectrum and affective disorders with a higher likelihood of exceeding normative percentile-based plasma NfL thresholds supports considering psychiatric diagnoses when interpreting blood NfL levels in neurological practice. In contrast, the limited associations between psychiatric diagnoses and plasma GFAP suggest that this marker may provide greater specificity for neurological conditions.

Adjustment for cardiometabolic burden and substance use modified associations between plasma NfL and GFAP and both psychiatric and neurological diagnoses, supporting their roles as modifiable factors influencing brain health. However, associations between plasma NfL and affective disorders persisted after adjustment for these factors as well as neurological comorbidities. Affective disorders may therefore influence circulating markers of neuroaxonal integrity beyond lifestyle factors and comorbidity. Though implementation of blood NfL in psychiatry requires further mechanistic investigation and careful consideration of relevant confounders, the associations we identified between this biomarker and affective disorders demonstrate its potential clinical utility as an accessible, biologically informed tool for psychiatry.

### Limitations

This study has several limitations. The UK Biobank sample consists of predominantly older White adults, restricting generalizability to younger, clinical, or more diverse populations. Biomarkers were measured cross-sectionally via proximity extension assay, reflecting relative rather than absolute abundance. Nevertheless, blood NfL levels measured via proximity extension assay and gold-standard immunoassays are strongly correlated.^31^ Another limitation arises from the definition of psychiatric and neurological diagnostic categories from lifetime diagnosis incidence, which may underestimate associations compared with current or more precisely defined clinical diagnoses. Detailed characterization of symptom severity, illness duration, and treatment exposure could clarify psychiatric state versus trait associations with blood NfL and GFAP and their relationship to treatment outcomes. Longitudinal studies are needed to determine whether blood NfL and GFAP track clinical course or treatment response. Finally, our study only allows associative investigation of biomarker alterations, precluding any causal or mechanistic inference. Additionally, blood NfL reflects both central and peripheral neuroaxonal injury. Studies integrating blood NfL and GFAP with neuroimaging and detailed phenotyping to clarify their relationship to brain pathology in psychiatric disorders will be essential to clarify the clinical and biological significance of these findings.

## Conclusions

This population-based study shows that plasma NfL is elevated in bipolar and depressive disorders and shows increased variability in schizophrenia spectrum, bipolar and depressive disorders, supporting its potential role as a biomarker in psychiatry. These findings also highlight the importance of considering psychiatric diagnoses when interpreting blood NfL levels in neurological practice.

## Data Availability

This research was conducted using the UK Biobank resource under application number [853656]. The data are available to researchers fulfilling the eligibility criteria through the UK Biobank access process.

## Acknowledgments and Conflict of Interest

The authors have no conflict of interest to declare. We gratefully acknowledge the participants of the UK Biobank for their invaluable contribution to this research. We also thank Barbora Rehák-Bucková for meaningful scientific exchange on normative modeling. FB was funded by Swiss National Science Foundation (Grant Nr. 217637) and EMDO Stiftung. The funders had no role in study design, data collection and analysis, decision to publish, or preparation of the manuscript.

## Author Contributions

Concept and design: AMJ, FB, and BBQ. Acquisition, analysis, or interpretation of data: AMJ and FB. Drafting of the manuscript: AMJ and FB. Critical review of the manuscript for important intellectual content: BBQ. Statistical analysis: AMJ. Obtained funding: BBQ and FB. Supervision: BBQ and FB.

