## Supplemental Tables for "Plasma Neurofilament Light Chain and Glial Fibrillary Acidic Protein in Psychiatric Disorders: A Large-Scale Normative Modeling Study"

| **UK Biobank variable label** | **UK Biobank variable** |
| --- | --- |
| NEFL;Neurofilament light polypeptide | olink_instance_0$nefl |
| GFAP;Glial fibrillary acidic protein | olink_instance_0$gfap |
| Age attended assessment centre | p21003_i0 |
| Sex | p31 |
| Body mass index (BMI) \| Instance 0 | p21001_i0 |
| Smoking status \| Instance 0 | p20116_i0 |
| Alcohol intake frequency \| Instance 0 | p1558_i0 |
| Date of attending assessment centre \| Instance 0 | p53_i0 |
| Townsend deprivation index at recruitment | p22189 |
| Qualifications \| Instance 0 | p6138_i0 |
| Ethnic background \| Instance 0 | p21000_i0 |
| Standard PRS for alzheimers disease (AD) | p26206 |
| Standard PRS for bipolar disorder (BD) | p26214 |
| Standard PRS for parkinsons disease (PD) | p26260 |
| Standard PRS for schizophrenia (SCZ) | p26275 |

***Supplementary Table 1. All UK Biobank Variables***

***Supplementary Table 2. Neurological and Psychiatric ICD10 Codes***

| **Diagnostic category** | **UK Biobank variable label (ICD10 code)** | **Number of entries** | **UK Biobank variable** |
| --- | --- | --- | --- |
| *Schizophrenia spectrum disorders* | Date F20 first reported (schizophrenia) | 147 | p130874 |
|  | Date F21 first reported (schizotypal disorder) | 3 | p130876 |
|  | Date F22 first reported (persistent delusional disorder) | 86 | p130878 |
|  | Date F23 first reported (acute and transient psychotic disorders) | 39 | p130880 |
|  | Date F25 first reported (schizoaffective disorders) | 32 | p130884 |
|  | Date F28 first reported (other nonorganic psychotic disorders) | 4 | p130886 |
|  | Date F29 first reported (unspecified nonorganic psychosis) | 74 | p130888 |
| *Bipolar disorder* | Date F30 first reported (manic episode) | 71 | p130890 |
|  | Date F31 first reported (bipolar affective disorder) | 259 | p130892 |
| *Recurrent depressive disorder* | Date F33 first reported (recurrent depressive disorder) | 326 | p130896 |
| *Depressive episode* | Date F32 first reported (depressive episode) | 5994 | p130894 |
| *Anxiety disorders* | Date F40 first reported (phobic anxiety disorders) | 460 | p130904 |
|  | Date F41 first reported (other anxiety disorders) | 4417 | p130906 |
| *Stress-related disorders* | Date F43 first reported (reaction to severe stress and adjustment disorders) | 1496 | p130910 |
| *Other psychiatric disorders* | Date F34 first reported (persistent mood [affective] disorders) | 91 | p130898 |
|  | Date F38 first reported (other mood [affective] disorders) | 20 | p130900 |
|  | Date F39 first reported (unspecified mood [affective] disorder) | 99 | p130902 |
|  | Date F42 first reported (obsessive compulsive disorder) | 90 | p130908 |
|  | Date F44 first reported (dissociative (conversion) disorders) | 49 | p130912 |
|  | Date F45 first reported (somatoform disorders) | 338 | p130914 |
|  | Date F48 first reported (other neurotic disorders) | 124 | p130916 |
|  | Date F99 first reported (mental disorder not otherwise specified) | 39 | p130990 |
| *Neurodegenerative disorders* | Date F00 first reported (dementia in Alzheimer's disease) | 517 | p130836 |
|  | Date F01 first reported (vascular dementia) | 296 | p130838 |
|  | Date F02 first reported (dementia in other diseases classified elsewhere) | 241 | p130840 |
|  | Date F03 first reported (unspecified dementia) | 860 | p130842 |
|  | Date G10 first reported (Huntington's disease) | 10 | p131012 |
|  | Date G12 first reported (spinal muscular atrophy and related syndromes) | 328 | p131016 |
|  | Date G13 first reported (systemic atrophies primarily affecting central nervous system in diseases classified elsewhere) | 6 | p131018 |
|  | Date G20 first reported (Parkinson's disease) | 877 | p131022 |
|  | Date G23 first reported (other degenerative diseases of basal ganglia) | 59 | p131028 |
|  | Date G25 first reported (other extrapyramidal and movement disorders) | 671 | p131032 |
|  | Date G30 first reported (Alzheimer's disease) | 718 | p131036 |
|  | Date G31 first reported (other degenerative diseases of nervous system not elsewhere classified) | 547 | p131038 |
|  | Date G32 first reported (other degenerative disorders of nervous system in diseases classified elsewhere) | 8 | p131040 |
| *Neuroinflammatory disorders* | Date G09 first reported (sequelae of inflammatory diseases of central nervous system) | 5 | p131010 |
|  | Date G35 first reported (multiple sclerosis) | 430 | p131042 |
|  | Date G36 first reported (other acute disseminated demyelination) | 9 | p131044 |
|  | Date G37 first reported (other demyelinating diseases of central nervous system) | 111 | p131046 |
| *Neuromuscular disorders* | Date G11 first reported (hereditary ataxia) | 50 | p131014 |
|  | Date G70 first reported (myasthenia gravis and other myoneural disorders) | 67 | p131092 |
|  | Date G71 first reported (primary disorders of muscles) | 39 | p131094 |
|  | Date G72 first reported (other myopathies) | 61 | p131096 |
|  | Date G73 first reported (disorders of myoneural junction and muscle in diseases classified elsewhere) | 6 | p131098 |
| *Cerebrovascular disorders* | Date G45 first reported (transient cerebral ischaemic attacks and related syndromes) | 1123 | p131056 |
|  | Date G46 first reported (vascular syndromes of brain in cerebrovascular diseases) | 35 | p131058 |
|  | Date I60 first reported (subarachnoid haemorrhage) | 224 | p131360 |
|  | Date I61 first reported (intracerebral haemorrhage) | 296 | p131362 |
|  | Date I62 first reported (other nontraumatic intracranial haemorrhage) | 171 | p131364 |
|  | Date I63 first reported (cerebral infarction) | 1191 | p131366 |
|  | Date I64 first reported (stroke not specified as haemorrhage or infarction) | 1135 | p131368 |
|  | Date I65 first reported (occlusion and stenosis of precerebral arteries not resulting in cerebral infarction) | 364 | p131370 |
|  | Date I66 first reported (occlusion and stenosis of cerebral arteries not resulting in cerebral infarction) | 28 | p131372 |
|  | Date I67 first reported (other cerebrovascular diseases) | 1510 | p131374 |
|  | Date I68 first reported (cerebrovascular disorders in diseases classified elsewhere) | 32 | p131376 |
|  | Date I69 first reported (sequelae of cerebrovascular disease) | 426 | p131378 |
| *Peripheral neuropathic disorders* | Date G60 first reported (hereditary and idiopathic neuropathy) | 72 | p131082 |
|  | Date G61 first reported (inflammatory polyneuropathy) | 71 | p131084 |
|  | Date G62 first reported (other polyneuropathies) | 654 | p131086 |
|  | Date G63 first reported (polyneuropathy in diseases classified elsewhere) | 198 | p131088 |
|  | Date G64 first reported (other disorders of peripheral nervous system) | 29 | p131090 |
| *Epilepsy* | Date G40 first reported (epilepsy) | 927 | p131048 |
|  | Date G41 first reported (status epilepticus) | 43 | p131050 |
| *Other neurological disorders* | Date G80 first reported (infantile cerebral palsy) | 27 | p131100 |
|  | Date G81 first reported (hemiplegia) | 563 | p131102 |
|  | Date G82 first reported (paraplegia and tetraplegia) | 67 | p131104 |
|  | Date G83 first reported (other paralytic syndromes) | 187 | p131106 |
|  | Date G91 first reported (hydrocephalus) | 143 | p131110 |
|  | Date G92 first reported (toxic encephalopathy) | 2 | p131112 |
|  | Date G93 first reported (other disorders of brain) | 781 | p131114 |
|  | Date G94 first reported (other disorders of brain in diseases classified elsewhere) | 27 | p131116 |
|  | Date G95 first reported (other diseases of spinal cord) | 232 | p131118 |
|  | Date G96 first reported (other disorders of central nervous system) | 68 | p131120 |
|  | Date G97 first reported (postprocedural disorders of nervous system not elsewhere classified) | 48 | p131122 |
|  | Date G98 first reported (other disorders of nervous system not elsewhere classified) | 238 | p131124 |
|  | Date G99 first reported (other disorders of nervous systems in diseases classified elsewhere) | 309 | p131126 |

***Supplementary Table 3. Normative Model Diagnostic Statistics***

| **Marker** | **MSLL** | **R^2^** | **Rho** | **SMSE** | **ShapiroW** |
| --- | --- | --- | --- | --- | --- |
| NfL | -20.472 | 0.303 | 0.556 | 0.697 | 0.994 |
| GFAP | -42.455 | 0.158 | 0.405 | 0.842 | 0.995 |
| *Abbreviations: MSLL: mean standardized log loss (model predictive density); R^2^: coefficient of determination (proportion of variance explained by the model)*  *Rho: correlation coefficient between observed and predicted values; SMSE: standardized mean squared error (SMSE<1.0 means performing better than simple mean model); ShapiroW: Shapiro-Wilks test for normality of model residuals* | | | | | |

***Supplementary Table 4. Cardiometabolic Burden ICD10 Codes***

| **Cardiometabolic category** | **UK Biobank variable label (ICD10 code)** | **Number of entries** | **UK Biobank variable** |
| --- | --- | --- | --- |
| *Heart disease* | Date I01 first reported (rheumatic fever with heart involvement) | 1 | p131272 |
|  | Date I05 first reported (rheumatic mitral valve diseases) | 113 | p131276 |
|  | Date I06 first reported (rheumatic aortic valve diseases) | 16 | p131278 |
|  | Date I07 first reported (rheumatic tricuspid valve diseases) | 168 | p131280 |
|  | Date I08 first reported (multiple valve diseases) | 714 | p131282 |
|  | Date I09 first reported (other rheumatic heart diseases) | 22 | p131284 |
|  | Date I20 first reported (angina pectoris) | 2434 | p131296 |
|  | Date I21 first reported (acute myocardial infarction) | 1575 | p131298 |
|  | Date I22 first reported (subsequent myocardial infarction) | 65 | p131300 |
|  | Date I23 first reported (certain current complications following acute myocardial infarction) | 12 | p131302 |
|  | Date I24 first reported (other acute ischaemic heart diseases) | 354 | p131304 |
|  | Date I25 first reported (chronic ischaemic heart disease) | 3455 | p131306 |
|  | Date I26 first reported (pulmonary embolism) | 883 | p131308 |
|  | Date I27 first reported (other pulmonary heart diseases) | 498 | p131310 |
|  | Date I30 first reported (acute pericarditis) | 50 | p131314 |
|  | Date I31 first reported (other diseases of pericardium) | 272 | p131316 |
|  | Date I33 first reported (acute and subacute endocarditis) | 61 | p131320 |
|  | Date I34 first reported (nonrheumatic mitral valve disorders) | 650 | p131322 |
|  | Date I35 first reported (nonrheumatic aortic valve disorders) | 642 | p131324 |
|  | Date I36 first reported (nonrheumatic tricuspid valve disorders) | 62 | p131326 |
|  | Date I37 first reported (pulmonary valve disorders) | 68 | p131328 |
|  | Date I38 first reported (endocarditis valve unspecified) | 100 | p131330 |
|  | Date I39 first reported (endocarditis and heart valve disorders in diseases classified elsewhere) | 2 | p131332 |
|  | Date I40 first reported (acute myocarditis) | 5 | p131334 |
|  | Date I41 first reported (myocarditis in diseases classified elsewhere) | 0 | p131336 |
|  | Date I42 first reported (cardiomyopathy) | 277 | p131338 |
|  | Date I43 first reported (cardiomyopathy in diseases classified elsewhere) | 11 | p131340 |
|  | Date I44 first reported (atrioventricular and left bundle branch block) | 972 | p131342 |
|  | Date I45 first reported (other conduction disorders) | 585 | p131344 |
|  | Date I46 first reported (cardiac arrest) | 301 | p131346 |
|  | Date I47 first reported (paroxysmal tachycardia) | 577 | p131348 |
|  | Date I48 first reported (atrial fibrillation and flutter) | 2737 | p131350 |
|  | Date I49 first reported (other cardiac arrhythmias) | 942 | p131352 |
|  | Date I50 first reported (heart failure) | 1731 | p131354 |
|  | Date I51 first reported (complications and ill-defined descriptions of heart disease) | 1751 | p131356 |
|  | Date I52 first reported (other heart disorders in diseases classified elsewhere) | 1 | p131358 |
|  | Date I70 first reported (atherosclerosis) | 311 | p131380 |
|  | Date I71 first reported (aortic aneurysm and dissection) | 353 | p131382 |
| *Hypertension* | Date I10 first reported (essential (primary) hypertension) | 10804 | p131286 |
|  | Date I11 first reported (hypertensive heart disease) | 84 | p131288 |
|  | Date I12 first reported (hypertensive renal disease) | 260 | p131290 |
|  | Date I13 first reported (hypertensive heart and renal disease) | 27 | p131292 |
|  | Date I15 first reported (secondary hypertension) | 29 | p131294 |
| *Diabetes* | Date E10 first reported (insulin dependent diabetes mellitus) | 397 | p130706 |
|  | Date E11 first reported (non-insulin dependent diabetes mellitus) | 2813 | p130708 |
|  | Date E13 first reported (other specified diabetes mellitus) | 45 | p130712 |
|  | Date E14 first reported (unspecified diabetes mellitus) | 1563 | p130714 |
| *Reduced renal function* | Date N17 first reported (acute renal failure) | 2112 | p132030 |
|  | Date N18 first reported (chronic renal failure) | 2388 | p132032 |
|  | Date N19 first reported (unspecified renal failure) | 339 | p132034 |

***Supplementary Table 5. Neurological Population Demographic and Clinical Characteristics***

| ***Characteristic*** | ***Reference population*** | ***Neuro-degenerative disorders*** | ***Neuro-inflammatory disorders*** | ***Neuro-muscular disorders*** | ***Cerebro-vascular disorders*** | ***Peripheral neuropathic disorders*** | ***Epilepsy*** | ***Other neurological disorders*** |
| --- | --- | --- | --- | --- | --- | --- | --- | --- |
| **Number of participants** | 5708 | 3109 | 489 | 213 | 3996 | 887 | 940 | 2284 |
| **Sex, N (%)** | NA | * | * | * | * | * | * |  |
| Male | 2628 (46.04) | 163 (52.75) | 141 (49.82) | 122 (37.42) | 2177 (36.32) | 1607 (33.99) | 531 (35.49) | 293 (35.95) |
| Female | 3080 (53.96) | 146 (47.25) | 142 (50.18) | 204 (62.58) | 3817 (63.68) | 3121 (66.01) | 965 (64.51) | 522 (64.05) |
| **Age, mean (SD)** | 56.26 (8.12) | 56.15 (8.39)* | 55.89 (8.0)* | 55.62 (8.56)* | 56.03 (8.16)* | 56.68 (8.11)* | 54.85 (8.07)* | 55.99 (7.97)* |
| **Body mass index, mean (SD)** | 27.14 (4.66 | 28.37 (5.38)* | 28.72 (5.69) | 28.74 (5.60)* | 28.25 (5.32)* | 27.73 (5.17)* | 27.86 (5.0)* | 27.81 (5.13)* |
| **Cardiometabolic burden, mean (SD)** | 0.73 (0.93) | 1.59 (1.25)* | 1.56 (1.23)* | 1.17 (1.22)* | 1.18 (1.16)* | 1.23 (1.14)* | 1.01 (1.09)* | 1.18 (1.16)* |
| **Tobacco smoking status, N (%)** | NA | * | * | * | * | * | * | * |
| Never | 3463 (60.67) | 133 (43.04) | 132 (46.64) | 164 (50.31) | 2830 (47.21) | 2361 (49.94) | 785 (52.47) | 415 (50.92) |
| Previous | 1935 (33.9) | 91 (29.45) | 93 (32.86) | 102 (31.29) | 2239 (37.35) | 1665 (35.22) | 525 (35.09) | 281 (34.48) |
| Current | 310 (5.43) | 85 (27.51) | 58 (20.49) | 60 (18.4) | 925 (15.43) | 702 (14.85) | 186 (12.43) | 119 (14.6) |
| **Alcohol consumption frequency, N (%)** | NA | * | * | * | * | * | * | * |
| Never | 387 (6.78) | 67 (21.68) | 59 (20.85) | 38 (11.66) | 700 (11.68) | 529 (11.19) | 153 (10.23) | 118 (14.48) |
| Special occasions only | 616 (10.79) | 47 (15.21) | 41 (14.49) | 47 (14.42) | 908 (15.15) | 690 (14.59) | 222 (14.84) | 93 (11.41) |
| 1-3x per month | 631 (11.05) | 37 (11.97) | 30 (10.6) | 42 (12.88) | 691 (11.53) | 541 (11.44) | 171 (11.43) | 92 (11.29) |
| 1-2x per week | 1588 (27.82) | 66 (21.36) | 55 (19.43) | 73 (22.39) | 1426 (23.79) | 1174 (24.83) | 369 (24.67) | 210 (25.77) |
| 3-4x per week | 1353 (23.7) | 46 (14.89) | 50 (17.67) | 72 (22.09) | 1150 (19.19) | 904 (19.12) | 298 (19.92) | 147 (18.04) |
| Daily or almost daily | 1133 (19.85) | 46 (14.89) | 48 (16.96) | 54 (16.56) | 1119 (18.67) | 890 (18.82) | 283 (18.92) | 155 (19.02) |
| **Highest educational qualification, N (%)** |  |  |  |  |  |  |  |  |
| College or university degree | 1952 (34.2) | 733 (23.58) | 161 (32.92) | 50 (23.47) | 986 (24.67) | 245 (27.62) | 234 (24.89) | 647 (28.33) |
| Associate degree or technical certification | 374 (6.55) | 225 (7.24) | 25 (5.11) | 18 (8.45) | 289 (7.23) | 60 (6.76) | 61 (6.49) | 170 (7.44) |
| Other professional licensure | 315 (5.52) | 193 (6.21) | 30 (6.13) | 13 (6.1) | 232 (5.81) | 48 (5.41) | 46 (4.89) | 129 (5.65) |
| Advanced high school completion | 628 (11) | 267 (8.59) | 74 (15.13) | 25 (11.74) | 327 (8.18) | 78 (8.79) | 86 (9.15) | 225 (9.85) |
| General high school completion | 1517 (26.58) | 724 (23.29) | 130 (26.58) | 57 (26.76) | 965 (24.15) | 229 (25.82) | 253 (26.91) | 565 (24.74) |
| None of the above | 831 (14.56) | 897 (28.85) | 64 (13.09) | 47 (22.07) | 1123 (28.1) | 203 (22.89) | 238 (25.32) | 502 (21.98) |
| Unknown | 91 (1.59) | 70 (2.25) | 5 (1.02) | 3 (1.41) | 74 (1.85) | 24 (2.71) | 22 (2.34) | 46 (2.01) |
| **Townsend deprivation index, mean (SD)** | -1.43 (3.05) | -1.16 (3.18) | -1.4 (3.14) | -0.32 (3.77) | -0.89 (3.27) | -0.58 (3.55) | -0.56 (3.43) | -0.8 (3.27) |
| **Ethnic background, N (%)** |  |  |  |  |  |  |  |  |
| White | 5295 (92.76) | 3001 (96.53) | 468 (95.71) | 204 (95.77) | 3784 (94.69) | 844 (95.15) | 886 (94.26) | 2132 (93.35) |
| Black | 106 (1.86) | 16 (0.51) | 1 (0.2) | 0 (0) | 34 (0.85) | 5 (0.56) | 12 (1.28) | 28 (1.23) |
| Asian | 124 (2.17) | 30 (0.96) | 6 (1.23) | 5 (2.35) | 74 (1.85) | 14 (1.58) | 16 (1.7) | 43 (1.88) |
| Mixed | 93 (1.63) | 32 (1.03) | 7 (1.43) | 3 (1.41) | 50 (1.25) | 7 (0.79) | 10 (1.06) | 42 (1.84) |
| Other | 73 (1.28) | 21 (0.68) | 4 (0.82) | 1 (0.47) | 41 (1.03) | 16 (1.8) | 12 (1.28) | 30 (1.31) |
| Unknown | 17 (0.3) | 9 (0.29) | 3 (0.61) | 0 (0) | 13 (0.33) | 1 (0.11) | 4 (0.43) | 9 (0.39) |

***Supplementary Table 6a. Demographic and Clinical Characteristic Comparative Statistics in Psychiatric Populations***

| **Characteristic** | **Stats** | **Schizophrenia spectrum disorders** | **Bipolar disorder** | **Recurrent depressive disorder** | **Depressive episode** | **Anxiety disorders** | **Stress-related disorders** | **Other psychiatric disorders** |
| --- | --- | --- | --- | --- | --- | --- | --- | --- |
| *Sex* | **χ2** | 5.04 | 1.40 | 8.89 | 113.76 | 155.27 | 53.11 | 28.95 |
|  | **p** | 0.02 | 0.24 | 0.003 | <0.001 | <0.001 | <0.001 | <0.001 |
| *Age* | **t** | -0.24 | -0.76 | -1.38 | -1.55 | 2.65 | -6.00 | -0.90 |
|  | **p** | 0.81 | 0.45 | 0.17 | 0.12 | 0.008 | 2.13E-09 | 0.37 |
| *Body mass index* | **t** | 4.49 | 5.49 | 5.93 | 11.91 | 6.10 | 5.21 | 3.78 |
|  | **p** | <0.001 | <0.001 | <0.001 | <0.001 | <0.001 | <0.001 | <0.001 |
| *Tobacco smoking status* | **χ2** | 234.60 | 108.53 | 90.79 | 385.31 | 291.15 | 98.41 | 102.66 |
|  | **p** | <0.001 | <0.001 | <0.001 | <0.001 | <0.001 | <0.001 | <0.001 |
| *Alcohol consumption frequency* | **χ2** | 109.64 | 87.34 | 20.36 | 167.17 | 122.74 | 47.91 | 66.18 |
|  | **p** | <0.001 | <0.001 | 0.001 | <0.001 | 8<0.001 | <0.001 | <0.001 |
| *Cardiometabolic burden* | **t** | 15.46 | 14.40 | 8.16 | 23.16 | 24.42 | 9.81 | 12.37 |
|  | **p** | <0.001 | <0.001 | <0.001 | <0.001 | <0.001 | <0.001 | <0.001 |
| *Highest educational qualification* | **χ2** | 24.18 | 12.461 | 15.34 | 108.84 | 109.09 | 10.15 | 26.00 |
|  | **p** | 0.002 | 0.132 | 0.05 | <0.001 | <0.001 | 0.25 | 0.001 |
| *Ethnic background* | **χ2** | 65.90 | 56.129 | 29.54 | 78.00 | 64.91 | 22.82 | 26.14 |
|  | **p** | <0.001 | <0.001 | 0.042 | <0.001 | <0.001 | 0.20 | 0.097 |
| *Townsend deprivation index* | **t** | 11.95 | 8.96 | 7.11 | 12.07 | 8.91 | 3.56 | 5.03 |
|  | **p** | <0.001 | <0.001 | <0.001 | <0.001 | <0.001 | <0.001 | <0.001 |

***Supplementary Table 6a. Demographic and Clinical Characteristic Comparative Statistics in Psychiatric Populations***

| **Characteristic** | **Stats** | **Neuro-**  **degenerative disorders** | **Neuro-**  **inflammatory disorders** | **Neuro-**  **muscular disorders** | **Cerebro-vascular disorders** | **Peripheral neuropathic disorders** | **Epilepsy** | **Other neurological disorders** |
| --- | --- | --- | --- | --- | --- | --- | --- | --- |
| *Sex* | **χ2** | 21.05 | 36.88 | 9.14 | 79.90 | 41.54 | 3.91 | 3.11 |
|  | **p** | <0.001 | <0.001 | 0.002 | <0.001 | <0.001 | 0.05 | 0.08 |
| *Age* | **t** | 38.97 | -2.97 | 3.83 | 34.20 | 11.22 | 4.85 | 11.69 |
|  | **p** | <0.001 | 0.003 | <0.001 | <0.001 | <0.001 | <0.001 | <0.001 |
| *Body mass index* | **t** | 5.84 | -0.54 | 5.64 | 10.46 | 12.64 | 5.59 | 8.58 |
|  | **p** | <0.001 | 0.59 | <0.001 | <0.001 | <0.001 | <0.001 | <0.001 |
| *Tobacco smoking status* | **χ2** | 117.20 | 108.24 | 18.12 | 319.85 | 108.54 | 108.88 | 198.16 |
|  | **p** | <0.001 | <0.001 | <0.001 | <0.001 | <0.001 | <0.001 | <0.001 |
| *Alcohol consumption frequency* | **χ2** | 150.27 | 51.67 | 16.22 | 141.48 | 88.91 | 78.45 | 130.53 |
|  | **p** | <0.001 | <0.001 | 0.006 | <0.001 | <0.001 | <0.001 | <0.001 |
| *Cardiometabolic burden* | **t** | 40.28 | 4.96 | 14.26 | 54.11 | 35.07 | 20.72 | 33.56 |
|  | **p** | <0.001 | <0.001 | <0.001 | <0.001 | <0.001 | <0.001 | <0.001 |
| *Highest educational qualification* | **χ2** | 326.23 | 11.17 | 16.54 | 315.72 | 54.00 | 86.45 | 79.43 |
|  | **p** | <0.001 | 0.19 | 0.035 | <0.001 | <0.001 | <0.001 | <0.001 |
| *Ethnic background* | **χ2** | 89.63 | 31.37 | 11.77 | 53.74 | 25.90 | 30.78 | 29.19 |
|  | **p** | <0.001 | 0.03 | 0.86 | <0.001 | 0.10 | 0.03 | 0.05 |
| *Townsend deprivation index* | **t** | 3.96 | 0.23 | 5.14 | 8.25 | 7.55 | 7.93 | 8.19 |
|  | **p** | <0.001 | 0.82 | <0.001 | <0.001 | <0.001 | <0.001 | <0.001 |

***Supplementary Table 7. Linear Model Covariates***

| **Marker** | **Covariate** | **Residual Cohen's d** | **95% CI (min)** | **95% CI (max)** | **p-value** | **Linear mixed effects model** |
| --- | --- | --- | --- | --- | --- | --- |
| *NfL* | Cardiometabolic burden | 0.123 | 0.105 | 0.141 | <0.001 | diagnoses + cardiometabolic burden |
|  | tobacco[T.Previous] | -0.046 | -0.085 | -0.007 | 0.021 | diagnoses + tobacco |
|  | tobacco[T.Current] | 0.013 | -0.048 | 0.073 | 0.684 |  |
|  | alcohol[T.Special occasions only] | 0.021 | -0.038 | 0.080 | 0.482 | diagnoses + alcohol |
|  | alcohol[T.One to three times a month] | -0.046 | -0.108 | 0.015 | 0.141 |  |
|  | alcohol[T.Once or twice a week] | -0.099 | -0.139 | -0.059 | <0.001 |  |
|  | alcohol[T.Three or four times a week] | -0.163 | -0.207 | -0.118 | <0.001 |  |
|  | alcohol[T.Daily or almost daily] | -0.161 | -0.207 | -0.116 | <0.001 |  |
|  | tobacco[T.Previous] | -0.058 | -0.099 | -0.016 | 0.006 | diagnoses + cardiometabolic burden + tobacco + alcohol |
|  | tobacco[T.Current] | 0.010 | -0.053 | 0.073 | 0.750 |  |
|  | alcohol[T.Special occasions only] | 0.030 | -0.049 | 0.109 | 0.461 |  |
|  | alcohol[T.One to three times a month] | -0.012 | -0.094 | 0.071 | 0.782 |  |
|  | alcohol[T.Once or twice a week] | -0.062 | -0.133 | 0.009 | 0.087 |  |
|  | alcohol[T.Three or four times a week] | -0.114 | -0.187 | -0.040 | 0.002 |  |
|  | alcohol[T.Daily or almost daily] | -0.120 | -0.194 | -0.046 | 0.001 |  |
|  | Cardiometabolic burden | 0.121 | 0.103 | 0.139 | <0.001 |  |
| *GFAP* | Cardiometabolic burden | -0.013 | -0.022 | -0.003 | 0.011 | diagnoses + cardiometabolic burden |
|  | tobacco[T.Previous] | -0.098 | -0.120 | -0.075 | <0.001 | diagnoses + tobacco |
|  | tobacco[T.Current] | -0.452 | -0.513 | -0.390 | <0.001 |  |
|  | alcohol[T.Special occasions only] | -0.022 | -0.100 | 0.056 | 0.581 | diagnoses + alcohol |
|  | alcohol[T.One to three times a month] | -0.043 | -0.125 | 0.039 | 0.309 |  |
|  | alcohol[T.Once or twice a week] | -0.124 | -0.193 | -0.055 | 0.0004 |  |
|  | alcohol[T.Three or four times a week] | -0.189 | -0.262 | -0.116 | <0.001 |  |
|  | alcohol[T.Daily or almost daily] | -0.252 | -0.317 | -0.187 | <0.001 |  |
|  | tobacco[T.Previous] | -0.069 | -0.110 | -0.027 | 0.0011 | diagnoses + cardiometabolic burden + tobacco + alcohol |
|  | tobacco[T.Current] | -0.438 | -0.501 | -0.375 | <0.001 |  |
|  | alcohol[T.Special occasions only] | -0.010 | -0.089 | 0.069 | 0.806 |  |
|  | alcohol[T.One to three times a month] | -0.041 | -0.124 | 0.041 | 0.328 |  |
|  | alcohol[T.Once or twice a week] | -0.124 | -0.195 | -0.053 | 0.0006 |  |
|  | alcohol[T.Three or four times a week] | -0.189 | -0.262 | -0.115 | <0.001 |  |
|  | alcohol[T.Daily or almost daily] | -0.227 | -0.302 | -0.153 | <0.001 |  |
|  | Cardiometabolic burden | -0.017 | -0.034 | 0.001 | 0.070 |  |

**Supplementary Table 8. NfL Linear Model Statistics**

| **Diagnostic category** | **Residual Cohen's d** | **95% CI (min)** | **95% CI (max)** | **FDR**  **p-value** | **Linear mixed effects model** |
| --- | --- | --- | --- | --- | --- |
| Schizophrenia spectrum disorders | -0.009 | -0.17 | 0.15 | 0.92 | diagnoses only |
| Bipolar disorder | 0.25 | 0.08 | 0.42 | 0.004 |  |
| Recurrent depressive disorder | 0.24 | 0.09 | 0.40 | 0.002 |  |
| Depressive episode | 0.08 | 0.04 | 0.12 | 0.0003 |  |
| Anxiety disorders | -0.05 | -0.09 | -0.003 | 0.04 |  |
| Stress-related disorders | 0.01 | -0.06 | 0.09 | 0.71 |  |
| Other psychiatric disorders | -0.04 | -0.14 | 0.06 | 0.46 |  |
| Neurodegenerative disorders | 0.34 | 0.28 | 0.39 | <0.001 |  |
| Neuroinflammatory disorders | 0.45 | 0.32 | 0.57 | <0.001 |  |
| Neuromuscular disorders | 0.25 | 0.06 | 0.44 | 0.01 |  |
| Cerebrovascular disorders | 0.17 | 0.12 | 0.22 | <0.001 |  |
| Peripheral neuropathic disorders | 0.64 | 0.55 | 0.73 | <0.001 |  |
| Epilepsy | 0.29 | 0.20 | 0.38 | <0.001 |  |
| Other neurological disorders | 0.13 | 0.07 | 0.19 | <0.001 |  |
| Schizophrenia spectrum disorders | -0.04 | -0.20 | 0.13 | 0.66 | diagnoses + cardiometabolic burden |
| Bipolar disorder | 0.21 | 0.04 | 0.38 | 0.02 |  |
| Recurrent depressive disorder | 0.23 | 0.08 | 0.39 | 0.006 |  |
| Depressive episode | 0.06 | 0.02 | 0.11 | 0.006 |  |
| Anxiety disorders | -0.07 | -0.11 | -0.02 | 0.006 |  |
| Stress-related disorders | 0.02 | -0.05 | 0.09 | 0.66 |  |
| Other psychiatric disorders | -0.04 | -0.14 | 0.06 | 0.48 |  |
| Neurodegenerative disorders | 0.28 | 0.22 | 0.33 | <0.001 |  |
| Neuroinflammatory disorders | 0.47 | 0.34 | 0.60 | <0.001 |  |
| Neuromuscular disorders | 0.21 | 0.02 | 0.41 | 0.04 |  |
| Cerebrovascular disorders | 0.07 | 0.02 | 0.13 | 0.009 |  |
| Peripheral neuropathic disorders | 0.54 | 0.44 | 0.63 | <0.001 |  |
| Epilepsy | 0.27 | 0.18 | 0.36 | <0.001 |  |
| Other neurological disorders | 0.10 | 0.04 | 0.16 | 0.006 |  |
| Schizophrenia spectrum disorders | -0.05 | -0.22 | 0.11 | 0.87 | diagnoses + tobacco |
| Bipolar disorder | 0.20 | 0.03 | 0.37 | 0.004 |  |
| Recurrent depressive disorder | 0.23 | 0.07 | 0.38 | 0.002 |  |
| Depressive episode | 0.06 | 0.02 | 0.10 | 0.0003 |  |
| Anxiety disorders | -0.07 | -0.12 | -0.02 | 0.034 |  |
| Stress-related disorders | 0.01 | -0.06 | 0.09 | 0.73 |  |
| Other psychiatric disorders | -0.04 | -0.14 | 0.06 | 0.45 |  |
| Neurodegenerative disorders | 0.28 | 0.22 | 0.33 | <0.001 |  |
| Neuroinflammatory disorders | 0.46 | 0.33 | 0.59 | <0.001 |  |
| Neuromuscular disorders | 0.21 | 0.02 | 0.40 | 0.01 |  |
| Cerebrovascular disorders | 0.07 | 0.02 | 0.13 | 0.000 |  |
| Peripheral neuropathic disorders | 0.54 | 0.44 | 0.63 | <0.001 |  |
| Epilepsy | 0.26 | 0.17 | 0.36 | <0.001 |  |
| Other neurological disorders | 0.09 | 0.03 | 0.16 | 0.000 |  |
| Schizophrenia spectrum disorders | -0.02 | -0.19 | 0.14 | 0.80 | diagnoses + alcohol |
| Bipolar disorder | 0.24 | 0.07 | 0.41 | 0.01 |  |
| Recurrent depressive disorder | 0.24 | 0.08 | 0.39 | 0.005 |  |
| Depressive episode | 0.07 | 0.03 | 0.11 | 0.003 |  |
| Anxiety disorders | -0.05 | -0.10 | -0.01 | 0.04 |  |
| Stress-related disorders | 0.01 | -0.06 | 0.08 | 0.80 |  |
| Other psychiatric disorders | -0.04 | -0.14 | 0.06 | 0.50 |  |
| Neurodegenerative disorders | 0.33 | 0.28 | 0.39 | <0.001 |  |
| Neuroinflammatory disorders | 0.44 | 0.31 | 0.57 | <0.001 |  |
| Neuromuscular disorders | 0.24 | 0.05 | 0.43 | 0.02 |  |
| Cerebrovascular disorders | 0.17 | 0.12 | 0.22 | <0.001 |  |
| Peripheral neuropathic disorders | 0.63 | 0.54 | 0.73 | <0.001 |  |
| Epilepsy | 0.28 | 0.19 | 0.37 | <0.001 |  |
| Other neurological disorders | 0.12 | 0.06 | 0.18 | 0.0003 |  |
| Schizophrenia spectrum disorders | -0.05 | -0.22 | 0.11 | 0.56 | diagnoses + cardiometabolic burden + tobacco + alcohol |
| Bipolar disorder | 0.20 | 0.03 | 0.37 | 0.03 |  |
| Recurrent depressive disorder | 0.23 | 0.07 | 0.38 | 0.009 |  |
| Depressive episode | 0.06 | 0.02 | 0.10 | 0.013 |  |
| Anxiety disorders | -0.07 | -0.12 | -0.02 | 0.008 |  |
| Stress-related disorders | 0.01 | -0.06 | 0.09 | 0.70 |  |
| Other psychiatric disorders | -0.04 | -0.14 | 0.06 | 0.45 |  |
| Neurodegenerative disorders | 0.28 | 0.22 | 0.33 | <0.001 |  |
| Neuroinflammatory disorders | 0.46 | 0.33 | 0.59 | <0.001 |  |
| Neuromuscular disorders | 0.21 | 0.02 | 0.40 | 0.04 |  |
| Cerebrovascular disorders | 0.07 | 0.02 | 0.13 | 0.01 |  |
| Peripheral neuropathic disorders | 0.54 | 0.44 | 0.63 | <0.001 |  |
| Epilepsy | 0.26 | 0.17 | 0.36 | <0.001 |  |
| Other neurological disorders | 0.09 | 0.03 | 0.16 | 0.008 |  |

***Supplementary Table 9. GFAP Linear Model Statistics***

| **Diagnostic category** | **Residual Cohen's d** | **95% CI (min)** | **95% CI (max)** | **FDR**  **p-value** | **Linear mixed effects model** |
| --- | --- | --- | --- | --- | --- |
| Schizophrenia spectrum disorders | -0.17 | -0.33 | -0.004 | 0.12 | diagnoses only |
| Bipolar disorder | 0.17 | -0.003 | 0.34 | 0.13 |  |
| Recurrent depressive disorder | 0.02 | -0.14 | 0.17 | 0.90 |  |
| Depressive episode | 0.01 | -0.02 | 0.04 | 0.66 |  |
| Anxiety disorders | 0.02 | -0.03 | 0.07 | 0.53 |  |
| Stress-related disorders | -0.03 | -0.11 | 0.04 | 0.53 |  |
| Other psychiatric disorders | -0.06 | -0.16 | 0.04 | 0.47 |  |
| Neurodegenerative disorders | 0.27 | 0.22 | 0.32 | <0.001 |  |
| Neuroinflammatory disorders | 0.35 | 0.22 | 0.48 | <0.001 |  |
| Neuromuscular disorders | -0.01 | -0.20 | 0.18 | 0.90 |  |
| Cerebrovascular disorders | -0.02 | -0.07 | 0.03 | 0.61 |  |
| Peripheral neuropathic disorders | -0.07 | -0.16 | 0.03 | 0.35 |  |
| Epilepsy | 0.17 | 0.08 | 0.26 | 0.0014 |  |
| Other neurological disorders | 0.06 | 0.007 | 0.12 | 0.10 |  |
| Schizophrenia spectrum disorders | -0.16 | -0.33 | -0.002 | 0.12 | diagnoses + cardiometabolic burden |
| Bipolar disorder | 0.17 | -0.001 | 0.34 | 0.12 |  |
| Recurrent depressive disorder | 0.02 | -0.14 | 0.17 | 0.89 |  |
| Depressive episode | 0.01 | -0.03 | 0.05 | 0.77 |  |
| Anxiety disorders | 0.02 | -0.02 | 0.07 | 0.46 |  |
| Stress-related disorders | -0.03 | -0.11 | 0.04 | 0.52 |  |
| Other psychiatric disorders | -0.06 | -0.15 | 0.04 | 0.46 |  |
| Neurodegenerative disorders | 0.28 | 0.22 | 0.33 | <0.001 |  |
| Neuroinflammatory disorders | 0.35 | 0.22 | 0.47 | <0.001 |  |
| Neuromuscular disorders | -0.008 | -0.20 | 0.18 | 0.93 |  |
| Cerebrovascular disorders | -0.007 | -0.06 | 0.04 | 0.89 |  |
| Peripheral neuropathic disorders | -0.06 | -0.15 | 0.04 | 0.46 |  |
| Epilepsy | 0.17 | 0.08 | 0.26 | 0.0017 |  |
| Other neurological disorders | 0.07 | 0.00 | 0.13 | 0.12 |  |
| Schizophrenia spectrum disorders | -0.13 | -0.29 | 0.03 | 0.31 | diagnoses + tobacco |
| Bipolar disorder | 0.19 | 0.02 | 0.36 | 0.06 |  |
| Recurrent depressive disorder | 0.04 | -0.11 | 0.20 | 0.62 |  |
| Depressive episode | 0.03 | -0.01 | 0.07 | 0.17 |  |
| Anxiety disorders | 0.03 | -0.01 | 0.08 | 0.21 |  |
| Stress-related disorders | -0.03 | -0.11 | 0.04 | 0.56 |  |
| Other psychiatric disorders | -0.05 | -0.15 | 0.05 | 0.46 |  |
| Neurodegenerative disorders | 0.27 | 0.22 | 0.32 | <0.001 |  |
| Neuroinflammatory disorders | 0.37 | 0.24 | 0.50 | <0.001 |  |
| Neuromuscular disorders | -0.01 | -0.20 | 0.18 | 0.92 |  |
| Cerebrovascular disorders | 0.02 | -0.04 | 0.07 | 0.88 |  |
| Peripheral neuropathic disorders | -0.05 | -0.14 | 0.05 | 0.46 |  |
| Epilepsy | 0.17 | 0.08 | 0.27 | 0.0007 |  |
| Other neurological disorders | 0.07 | 0.007 | 0.13 | 0.06 |  |
| Schizophrenia spectrum disorders | -0.19 | -0.35 | -0.03 | 0.08 | diagnoses + alcohol |
| Bipolar disorder | 0.15 | -0.02 | 0.32 | 0.20 |  |
| Recurrent depressive disorder | 0.007 | -0.15 | 0.16 | 0.98 |  |
| Depressive episode | -0.001 | -0.04 | 0.04 | 0.98 |  |
| Anxiety disorders | 0.02 | -0.03 | 0.06 | 0.62 |  |
| Stress-related disorders | -0.04 | -0.11 | 0.04 | 0.50 |  |
| Other psychiatric disorders | -0.06 | -0.16 | 0.04 | 0.41 |  |
| Neurodegenerative disorders | 0.27 | 0.21 | 0.32 | <0.001 |  |
| Neuroinflammatory disorders | 0.34 | 0.21 | 0.47 | <0.001 |  |
| Neuromuscular disorders | -0.02 | -0.21 | 0.17 | 0.98 |  |
| Cerebrovascular disorders | -0.02 | -0.07 | 0.03 | 0.62 |  |
| Peripheral neuropathic disorders | -0.07 | -0.17 | 0.02 | 0.26 |  |
| Epilepsy | 0.16 | 0.06 | 0.25 | 0.0042 |  |
| Other neurological disorders | 0.06 | -0.007 | 0.12 | 0.20 |  |
| Schizophrenia spectrum disorders | -0.13 | -0.29 | 0.03 | 0.28 | diagnoses + cardiometabolic burden + tobacco + alcohol |
| Bipolar disorder | 0.19 | 0.02 | 0.36 | 0.09 |  |
| Recurrent depressive disorder | 0.04 | -0.11 | 0.20 | 0.65 |  |
| Depressive episode | 0.03 | -0.01 | 0.07 | 0.33 |  |
| Anxiety disorders | 0.03 | -0.01 | 0.08 | 0.33 |  |
| Stress-related disorders | -0.03 | -0.11 | 0.04 | 0.46 |  |
| Other psychiatric disorders | -0.05 | -0.15 | 0.05 | 0.46 |  |
| Neurodegenerative disorders | 0.27 | 0.22 | 0.32 | <0.001 |  |
| Neuroinflammatory disorders | 0.37 | 0.24 | 0.50 | <0.001 |  |
| Neuromuscular disorders | -0.01 | -0.20 | 0.18 | 0.91 |  |
| Cerebrovascular disorders | 0.02 | -0.04 | 0.07 | 0.64 |  |
| Peripheral neuropathic disorders | -0.05 | -0.14 | 0.05 | 0.46 |  |
| Epilepsy | 0.17 | 0.08 | 0.27 | 0.0013 |  |
| Other neurological disorders | 0.07 | 0.007 | 0.13 | 0.09 |  |

***Supplementary Table 10. NfL and GFAP Percentiles and Odds Ratios***

| **Marker** | **Normative percentile** | **Diagnostic category** | **% above threshold** | **Adjusted odds ratio** | **95% CI (low)** | **95% CI (high)** |
| --- | --- | --- | --- | --- | --- | --- |
| *NfL* | 80th | Reference population | 19.25 | NA | NA | NA |
|  |  | Schizophrenia spectrum disorders | 27.18 | 0.92 | 0.70 | 1.20 |
|  |  | Bipolar disorder | 30.39 | 1.33 | 1.01 | 1.74 |
|  |  | Recurrent depressive disorder | 29.45 | 1.47 | 1.15 | 1.89 |
|  |  | Depressive episode | 24.91 | 1.14 | 1.06 | 1.23 |
|  |  | Anxiety disorders | 23.12 | 0.93 | 0.86 | 1.01 |
|  |  | Stress-related disorders | 23.20 | 1.05 | 0.92 | 1.19 |
|  |  | Other psychiatric disorders | 22.09 | 0.88 | 0.74 | 1.05 |
|  |  | Neurodegenerative disorders | 31.59 | 1.40 | 1.28 | 1.52 |
|  |  | Neuroinflammatory disorders | 33.13 | 1.62 | 1.33 | 1.97 |
|  |  | Neuromuscular disorders | 34.74 | 1.31 | 0.98 | 1.76 |
|  |  | Cerebrovascular disorders | 28.15 | 1.07 | 0.99 | 1.17 |
|  |  | Peripheral neuropathic disorders | 40.92 | 1.97 | 1.71 | 2.27 |
|  |  | Epilepsy | 34.36 | 1.55 | 1.35 | 1.79 |
|  |  | Other neurological disorders | 29.68 | 1.19 | 1.07 | 1.31 |
|  | 90th | Reference population | 8.06 | NA | NA | NA |
|  |  | Schizophrenia spectrum disorders | 16.18 | 1.10 | 0.79 | 1.53 |
|  |  | Bipolar disorder | 18.02 | 1.59 | 1.14 | 2.21 |
|  |  | Recurrent depressive disorder | 14.11 | 1.48 | 1.07 | 2.05 |
|  |  | Depressive episode | 11.80 | 1.17 | 1.06 | 1.29 |
|  |  | Anxiety disorders | 10.32 | 0.86 | 0.77 | 0.96 |
|  |  | Stress-related disorders | 10.36 | 1.02 | 0.85 | 1.21 |
|  |  | Other psychiatric disorders | 10.43 | 0.90 | 0.71 | 1.14 |
|  |  | Neurodegenerative disorders | 15.92 | 1.36 | 1.22 | 1.52 |
|  |  | Neuroinflammatory disorders | 18.61 | 1.89 | 1.49 | 2.40 |
|  |  | Neuromuscular disorders | 21.13 | 1.53 | 1.08 | 2.16 |
|  |  | Cerebrovascular disorders | 14.29 | 1.09 | 0.97 | 1.22 |
|  |  | Peripheral neuropathic disorders | 25.14 | 2.29 | 1.95 | 2.71 |
|  |  | Epilepsy | 18.19 | 1.60 | 1.34 | 1.91 |
|  |  | Other neurological disorders | 15.19 | 1.19 | 1.05 | 1.36 |
|  | 95th | Reference population | 3.75 | NA | NA | NA |
|  |  | Schizophrenia spectrum disorders | 11.65 | 1.68 | 1.14 | 2.46 |
|  |  | Bipolar disorder | 9.54 | 1.44 | 0.93 | 2.24 |
|  |  | Recurrent depressive disorder | 7.67 | 1.62 | 1.06 | 2.48 |
|  |  | Depressive episode | 5.97 | 1.21 | 1.06 | 1.39 |
|  |  | Anxiety disorders | 4.76 | 0.77 | 0.66 | 0.90 |
|  |  | Stress-related disorders | 4.75 | 0.95 | 0.74 | 1.22 |
|  |  | Other psychiatric disorders | 4.79 | 0.83 | 0.59 | 1.16 |
|  |  | Neurodegenerative disorders | 7.88 | 1.23 | 1.06 | 1.43 |
|  |  | Neuroinflammatory disorders | 11.86 | 2.36 | 1.77 | 3.15 |
|  |  | Neuromuscular disorders | 14.08 | 1.89 | 1.26 | 2.85 |
|  |  | Cerebrovascular disorders | 7.18 | 0.98 | 0.84 | 1.14 |
|  |  | Peripheral neuropathic disorders | 14.66 | 2.32 | 1.89 | 2.86 |
|  |  | Epilepsy | 9.36 | 1.54 | 1.22 | 1.95 |
|  |  | Other neurological disorders | 9.02 | 1.42 | 1.21 | 1.68 |
| *GFAP* | 80th | Reference population | 19.25 | NA | NA | NA |
|  |  | Schizophrenia spectrum disorders | 27.18 | 0.73 | 0.54 | 0.98 |
|  |  | Bipolar disorder | 30.39 | 1.23 | 0.93 | 1.63 |
|  |  | Recurrent depressive disorder | 29.45 | 0.99 | 0.75 | 1.30 |
|  |  | Depressive episode | 24.91 | 1.00 | 0.93 | 1.08 |
|  |  | Anxiety disorders | 23.12 | 1.08 | 1.00 | 1.17 |
|  |  | Stress-related disorders | 23.20 | 1.01 | 0.89 | 1.15 |
|  |  | Other psychiatric disorders | 22.09 | 0.84 | 0.70 | 1.00 |
|  |  | Neurodegenerative disorders | 31.59 | 1.63 | 1.49 | 1.78 |
|  |  | Neuroinflammatory disorders | 33.13 | 1.58 | 1.29 | 1.92 |
|  |  | Neuromuscular disorders | 34.74 | 1.00 | 0.73 | 1.38 |
|  |  | Cerebrovascular disorders | 28.15 | 1.02 | 0.93 | 1.11 |
|  |  | Peripheral neuropathic disorders | 40.92 | 0.90 | 0.76 | 1.07 |
|  |  | Epilepsy | 34.36 | 1.29 | 1.11 | 1.50 |
|  |  | Other neurological disorders | 29.68 | 1.03 | 0.93 | 1.15 |
|  | 90th | Reference population | 8.06 | NA | NA | NA |
|  |  | Schizophrenia spectrum disorders | 16.18 | 0.80 | 0.54 | 1.20 |
|  |  | Bipolar disorder | 18.02 | 1.07 | 0.72 | 1.58 |
|  |  | Recurrent depressive disorder | 14.11 | 1.26 | 0.89 | 1.78 |
|  |  | Depressive episode | 11.80 | 1.00 | 0.90 | 1.10 |
|  |  | Anxiety disorders | 10.32 | 1.10 | 0.98 | 1.22 |
|  |  | Stress-related disorders | 10.36 | 0.94 | 0.79 | 1.13 |
|  |  | Other psychiatric disorders | 10.43 | 0.85 | 0.66 | 1.09 |
|  |  | Neurodegenerative disorders | 15.92 | 1.91 | 1.71 | 2.13 |
|  |  | Neuroinflammatory disorders | 18.61 | 1.78 | 1.38 | 2.28 |
|  |  | Neuromuscular disorders | 21.13 | 1.02 | 0.67 | 1.57 |
|  |  | Cerebrovascular disorders | 14.29 | 1.02 | 0.90 | 1.15 |
|  |  | Peripheral neuropathic disorders | 25.14 | 0.88 | 0.69 | 1.11 |
|  |  | Epilepsy | 18.19 | 1.42 | 1.17 | 1.72 |
|  |  | Other neurological disorders | 15.19 | 1.07 | 0.92 | 1.23 |
|  | 95th | Reference population | 3.75 | NA | NA | NA |
|  |  | Schizophrenia spectrum disorders | 11.65 | 0.82 | 0.47 | 1.44 |
|  |  | Bipolar disorder | 9.54 | 1.52 | 0.92 | 2.53 |
|  |  | Recurrent depressive disorder | 7.67 | 1.49 | 0.92 | 2.40 |
|  |  | Depressive episode | 5.97 | 1.13 | 0.98 | 1.32 |
|  |  | Anxiety disorders | 4.76 | 1.06 | 0.90 | 1.25 |
|  |  | Stress-related disorders | 4.75 | 1.00 | 0.77 | 1.31 |
|  |  | Other psychiatric disorders | 4.79 | 0.93 | 0.66 | 1.32 |
|  |  | Neurodegenerative disorders | 7.88 | 1.89 | 1.61 | 2.22 |
|  |  | Neuroinflammatory disorders | 11.86 | 1.87 | 1.32 | 2.63 |
|  |  | Neuromuscular disorders | 14.08 | 1.03 | 0.55 | 1.91 |
|  |  | Cerebrovascular disorders | 7.18 | 0.90 | 0.75 | 1.08 |
|  |  | Peripheral neuropathic disorders | 14.66 | 0.89 | 0.63 | 1.27 |
|  |  | Epilepsy | 9.36 | 1.88 | 1.46 | 2.42 |
|  |  | Other neurological disorders | 9.02 | 1.24 | 1.02 | 1.52 |
