## Supplemental Figures for "Plasma Neurofilament Light Chain and Glial Fibrillary Acidic Protein in Psychiatric Disorders: A Large-Scale Normative Modeling Study"

**Supplementary Figures**

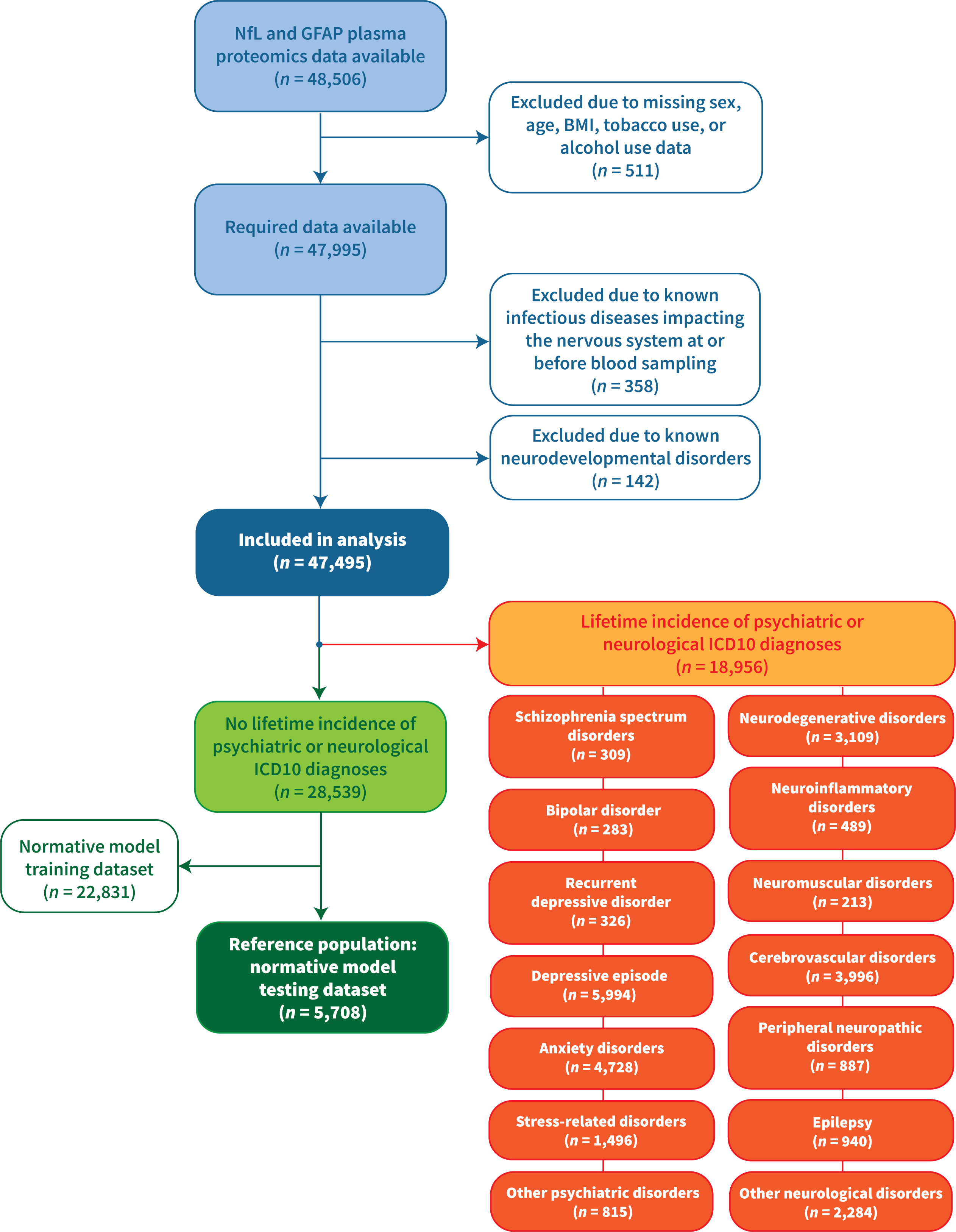

*Supplementary Figure 1. Consort Diagram*

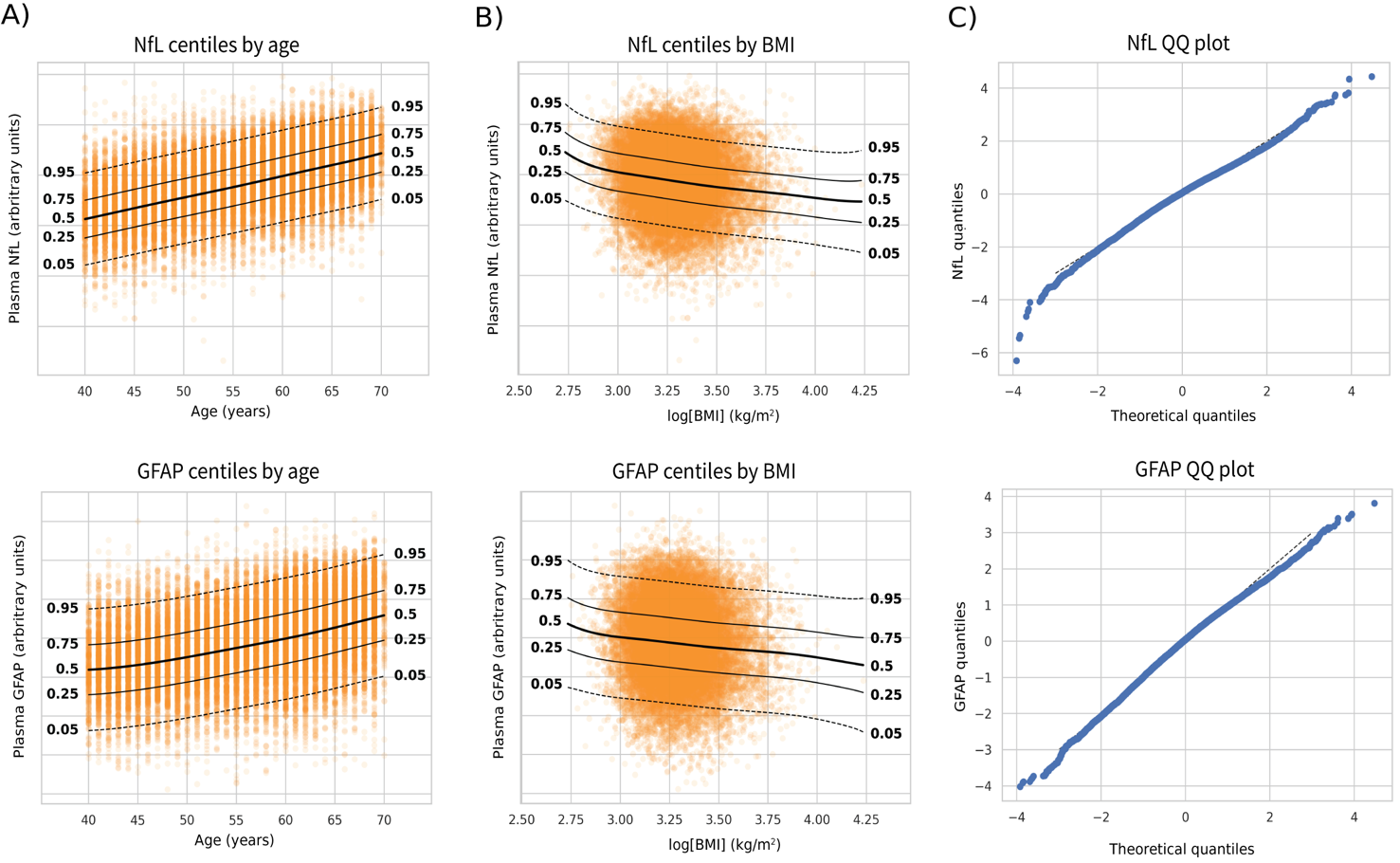

*Supplementary Figure 2. Normative Modeling of Plasma NfL and GFAP Levels*

A) Centiles of plasma NfL and GFAP levels based on age for the normative modeling training dataset. B) Centiles of plasma NfL and GFAP based on log-transformed body mass index (BMI) for the normative modeling training dataset. C) Quantile-quantile plots of NfL and GFAP normative model predictions versus actual plasma NfL and GFAP levels. Supplementary Table 3 lists diagnostic statistics for both models, which indicate robust and reliable predictive power.

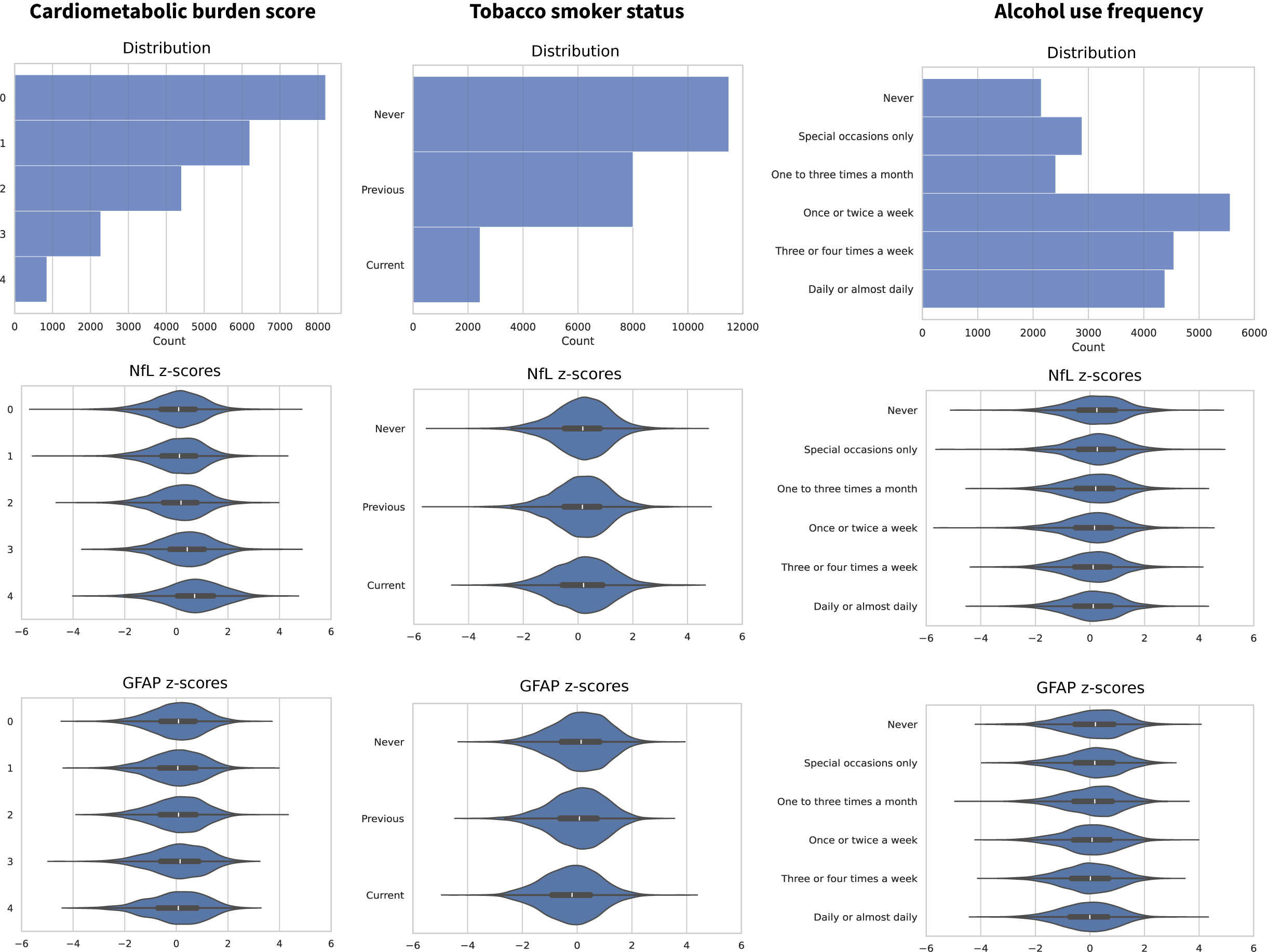

*Supplementary Figure 3. Clinical and Lifestyle Covariates*

Covariate distributions and associations with plasma NfL and GFAP z-scores based on age, sex, and BMI for reference, psychiatric, and neurological populations combined. Cardiometabolic burden scores were calculated by aggregating ICD10 codes in participant’s medical records related to heart disease, hypertension, diabetes, and reduced renal function (Supplementary Table 4), with one point added for each category with at least once diagnosis present. Self-report tobacco and alcohol use information was acquired from lifestyle questionnaires (Supplementary Table 1).

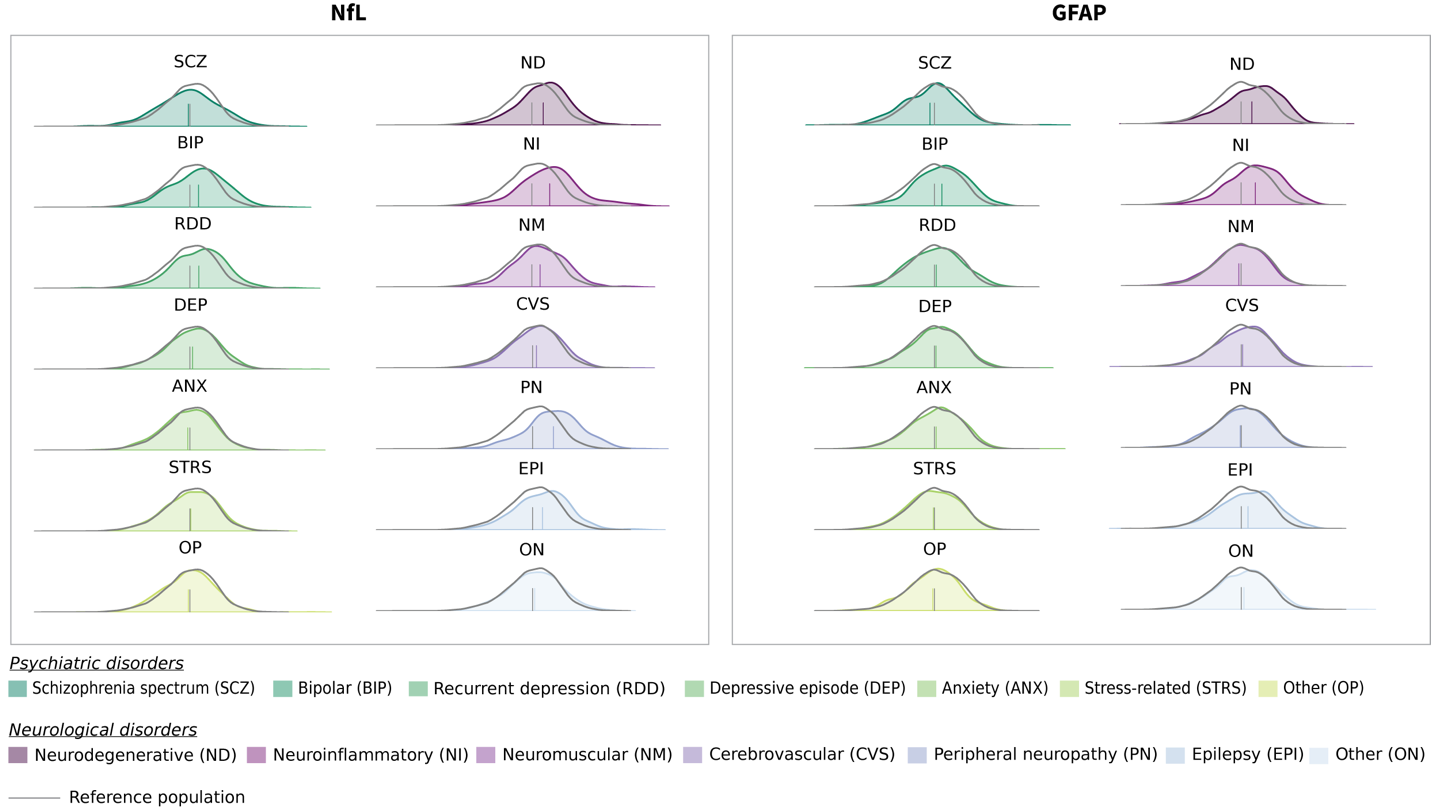

*Supplementary Figure 4. Adjusted NfL and GFAP Z-score Distributions*

Kernel density estimate plots (*seaborn* v0.13.2) of NfL and GFAP z-scores for each psychiatric and neurological diagnostic category compared to the reference population, adjusted for psychiatric and neurological comorbidities, cardiometabolic burden, tobacco use, and alcohol use. Vertical lines represent the mean for each distribution.
